# Unraveling Regional Variability in Dengue Outbreaks in Brazil: leveraging the Moving Epidemics Method (MEM) and Climate Data to Optimize Vector Control Strategies

**DOI:** 10.1101/2025.05.09.25327307

**Authors:** Ayrton Sena Gouveia, Marcelo Ferreira da Costa Gomes, Iasmim Ferreira de Almeida, Raquel Martins Lana, Leonardo Soares Bastos, Lucas Monteiro Bianchi, Sara de Souza Oliveira, Eduardo Correa Araujo⁴, Danielle Andreza da Cruz Ferreira, Dalila Machado Botelho Oliveira, Vinicius Barbosa Godinho, Luã Bida Vacaro, Thais Irene Souza Riback, Oswaldo Gonçalves Cruz, Flávio Codeço Coelho, Cláudia Torres Codeço

## Abstract

A country with continental dimensions like Brazil, characterized by heterogeneity of climates, biomes, natural resources, population density, socioeconomic conditions, and regional challenges, also exhibits significant spatial variation in dengue outbreaks. This study aimed to characterize Brazilian territory based on epidemiological and climate data to determine the optimal time to guide preventive and control strategies. To achieve this, the Moving Epidemics Method (MEM) was employed to analyze dengue historical patterns using 14-year disease data (2010–2023) aggregated by the 120 Brazilian Health Macro-Regions (HMR). Statistical outputs from MEM included the mean outbreak onset, duration, and variation of these measurements, pre– and post-epidemic thresholds, and the high-intensity level of cases. Environmental data used includes mean annual precipitation, temperature, and altitude, as well as the Köppen Climate Classification of each area. A multivariate cluster analysis using the k-means algorithm was applied to MEM outputs and climate data. Four clusters/regions were identified, with the mean temperature, mean precipitation, mean outbreak onset, high-intensity level of cases, and mean altitude explaining 80% of the centroid variation among the clusters. Region 1 (North-Northwest) encompasses areas with the highest temperatures, precipitation, and early outbreak onset, in February. Region 2a (Northeast) has the lowest precipitation and a later onset, in March. Region 3 (Southeast) presents higher altitude, and early outbreak onset in February; while Region 4 (South) has a lower temperature, with onset in March. To better adjust the results, the unique Roraima state HMR state was manually classified as Region 2b (Roraima) because of its outbreak onset in July and the highest precipitation volume. The results suggested preventive and control measures should be implemented first in Regions North-Northwest and Southeast, followed by Regions Northeast, South, and Roraima, highlighting the importance of regional vector control measures based on historical and climatic patterns. Integrating these findings with monitoring systems and fostering cross-sector collaboration can enhance surveillance and mitigate future outbreaks. The proposed methodology also holds potential for application in controlling other mosquito-transmitted viral diseases, expanding its public health impact.

**Author summary:** Dengue fever, a mosquito-transmitted viral disease, represents a significant public health challenge in tropical countries like Brazil. Transmission patterns vary widely across the country, shaped by diverse geography, climate, and local conditions. This study analyzed 14-year dengue data (2010–2023) from 120 Health Macro-Regions (HMR) in Brazil, integrating epidemiological and climate data to understand regional variations and optimize the timing of vector control strategies. This study applied the Moving Epidemics Method (MEM) output metrics, such as intensity and outbreak onset, along with environmental data (e.g., temperature and precipitation) and altitude in a multivariate cluster analysis to identify similar transmission seasons potentially driven by comparable climatic conditions. As a result, five regions emerged with similar outbreak patterns, emphasizing the need for tailored interventions. For example, some regions require vector control measures before February (∼10 weeks of uncertainty), while others can delay until June (∼13 weeks of uncertainty), highlighting the importance of locally adapted strategies. By integrating environmental data with traditional epidemiological approaches, this research offers valuable insights for enhancing disease control efforts. This methodology provides a framework for addressing other mosquito-borne diseases, including Zika and chikungunya.

## Introduction

Dengue is a viral disease transmitted by mosquitoes, with *Aedes aegypti* as the primary vector in tropical regions worldwide. Meanwhile, *Aedes albopictus* predominates in temperate and subtropical regions, such as Europe and East Asia. The disease is recognized as one of the top threats to global health (1) due to its high burden, epidemic proneness and increasing spatial spread. The frequency and intensity of dengue outbreaks in tropical regions have been intensified by rapid unplanned urbanization, growing population density, significant social inequalities, and increased mobility (2). More recently, the erosion of climate barriers is facilitating the swift spread of this disease to higher latitudes and altitudes. In endemic areas, dengue transmission exhibits a seasonal pattern, with increased incidence during periods of higher temperature, precipitation and humidity, delimiting transmission seasons (3–5).

Dengue seasonality is linked to fluctuations in mosquito abundance and vectorial capacity, mediated by temperature and precipitation. Warmer temperatures accelerate mosquito development, while higher precipitation increases the availability of breeding sites (6–9). These seasonal effects are further exacerbated by inadequate urban infrastructure, accumulation of garbage and unprotected water reservoirs. Other nonseasonal factors such as the population’s immune profile and the introduction of new strains or serotypes also play a crucial role in determining dengue incidence patterns (10–12). The complexity and variability of these interactions make preventing and controlling dengue outbreaks a permanent challenge.

In Brazil, significant dengue epidemics have occurred following the introduction of dengue serotypes, DENV1 in 1987, DENV2 in 1991, DENV3 in 2001 and DENV4 in 2012 (13). However, the serotype identification is only restricted to a small fraction of the reported cases (14). In 2024, dengue has set a record high in incidence and lethality, mostly caused by DENV1-2 (15,16). Brazil is a vast country, stretching from latitude 33°45’ South to 5°16’ North and from longitude 73°59’ West to 34°47’ East, encompassing equatorial, tropical, semi-arid and subtropical climates. Historically, dengue seasons are not synchronized across the country, starting earlier in the northern and central-western states, followed by the southeastern states and, at last, the northeastern states (17). This pattern suggests that the country could benefit from it by being able to focus control activities on one region at a time.

Many dengue prevention strategies are more efficient when implemented at the beginning of the season: breeding site removal efforts, education campaigns, and targeted insecticide treatments, among others. Once transmission has begun, surveillance and control efforts shift focus to health assistance. Defining typical onset and season duration can help more timely decision-making and resource allocation (18).

The moving epidemic method (MEM) is a well-established technique that was developed for flu-like illness (ILI) seasons. ILI, as dengue, is characterized by strong seasonality and spatial variability. The application of MEM to characterize typical onset, duration and activity levels of ILI seasons has been proven useful in Europe, Asia and Africa (19–22).

This work aims to extend the application of MEM to dengue by extracting typical seasonal parameters for this disease from all Health Macro-Regions (HMRs) of Brazil. Once collected, these seasonal metrics are harmonized with climate data and used in a multivariate cluster analysis, to identify areas with similar transmission seasons and comparable climatic conditions in Brazil. This approach seeks to generate a stratification of the country into areas with similar seasonality, which can directly inform disease control strategies and help to better determine the most opportune periods for implementing preventive educational and entomological interventions. It also enables health services to be prepared for potential increases in patients presenting symptoms of this disease. The same methodology can be applied to other countries and other strongly seasonal diseases.

## Methodology

### Study Area

Brazil is the largest country in Latin America, covering over 8.5 million km², with a population of more than 203 million (IBGE) (23). It is divided into five regions (North, Northeast, Southeast, South, and Center-West), 27 states, and 5,570 municipalities. Municipalities are further grouped into 120 HMRs. This stratification is designed to facilitate the organization of public health services and medical infrastructure, ensuring equitable access to healthcare for the population, as established by the Brazilian Ministry of Health (24). In this study, HMRs served as the primary units of analysis (Figure 1).

**Figure 1.**
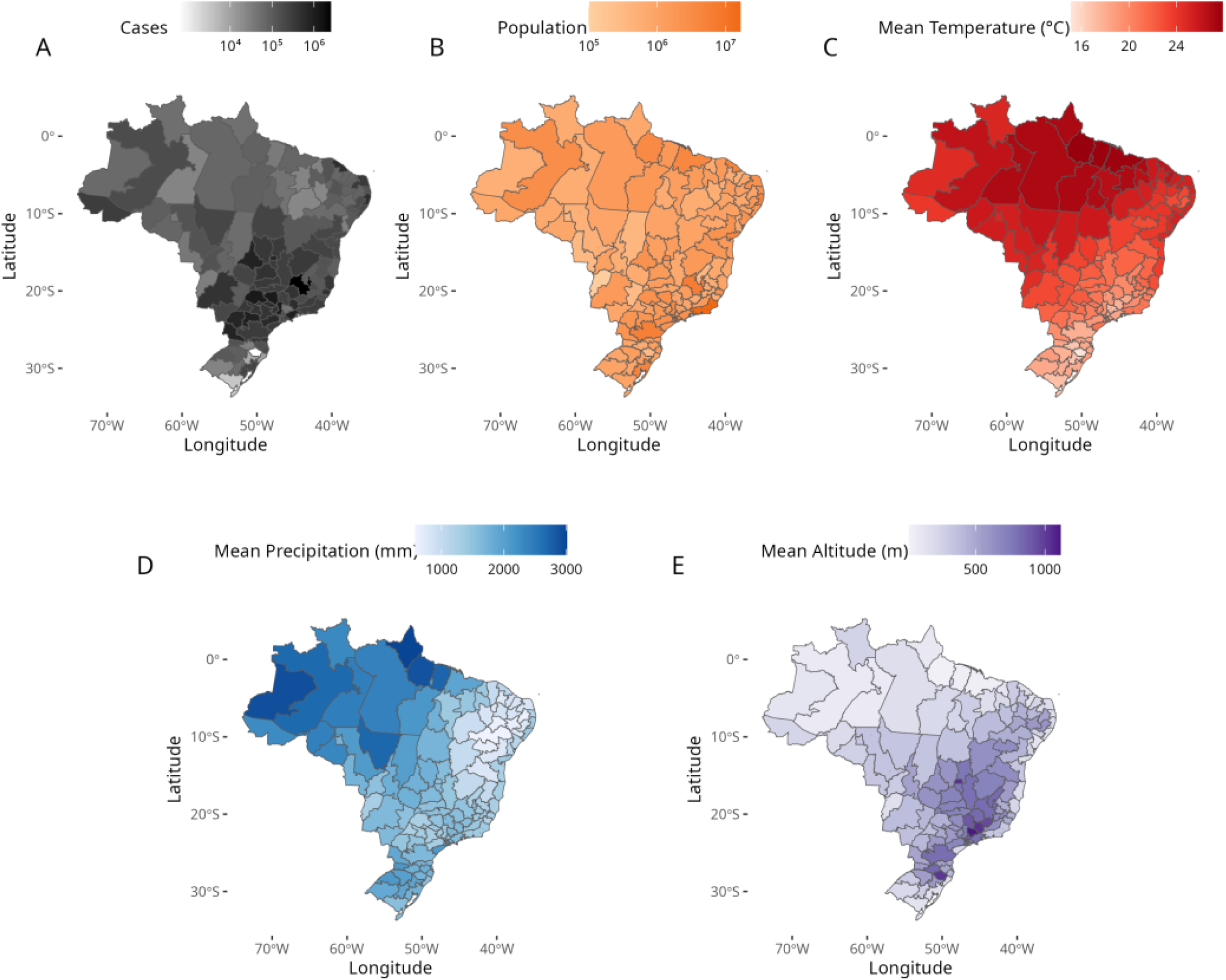
Distribution of dengue, population and climate among the 120 Health Macro Regions, Brazil. (A) Dengue cases reported from 2010 to 2023; (B) population size (2022); (C) mean annual temperature (1950–1990); (D) mean annual precipitation (1950–1990); (E) mean altitude. Service Layer Credits: Sources: https://www.ibge.gov.br/geociencias/organizacao-do-territorio/malhas-territoriais/15774-malhas.html?=&t=downloads

### Disease data

The dengue cases were obtained through the Infodengue platform (https://info.dengue.mat.br) (25), a Brazilian early warning system which harmonizes weather data and officially reported dengue cases by epiweek. The study used a dataset containing all reported dengue cases from January 1st 2010 to October 7th 2023 (epiweek 40 of 2023) for all 5,570 Brazilian municipalities, later aggregated in the 120 HMRs. The incidence rate per 100,000 inhabitants was calculated by week, and normalized by the population of each HMR for the corresponding year, obtained by aggregating the population data of municipalities, comprising each HMR, as provided by Instituto Brasileiro de Geografia e Estatística (IBGE) (23).

In Brazil, the surveillance of mosquito-transmitted viral diseases is primarily conducted through the suspected cases reported to the National Notification Database (SINAN). Dengue cases are characterized by fever between 38°C and 40°C, pruritus, rash, retro-orbital pain, and the absence of another apparent cause (26). Laboratory confirmation of cases is performed using various methods, such as real-time polymerase chain reaction (RT-PCR), which detects the presence of viral RNA, and serology, which checks for specific antibodies presence. When laboratory confirmation is not available, confirmation may be based on clinical-epidemiological criteria, including characteristic symptomatology and spatial proximity to confirmed cases (27).

### Environmental data

The country’s climate is classified according to the Koppen climate classification system. This system (28) uses precipitation, vegetation, temperature, latitude, longitude, and altitude for the classification of climate into 31 types, organized into 5 major categories: tropical (A), dry (B), humid subtropical (C), temperate continental (D), and polar (E).

Under this system, which was applied to Brazil in 2014, the country is divided into three climate types (29), tropical (with 4 subtypes: Af, Am, As and Aw), humid subtropical (with 4 subtypes: Cfa, Cfb, Cwa, Cwb), and dry (1 subtype: BSh) (Table 1 in supplementary information 1). Climate diversity is higher along the coast where dry, tropical, and subtropical climates predominate. The North region is predominantly tropical, without dry seasons and with monsoons. In contrast, the Center-West exhibits a dominance of a tropical climate with dry summers, and the Southern Region is predominantly subtropical. This climatic diversity directly influences vegetation distribution, hydrological cycles, and economic activities (agriculture and livestock). It also shapes regions with higher arbovirus risk, making climate classification a crucial factor for understanding regional dynamics in Brazil.

**Table 1.**
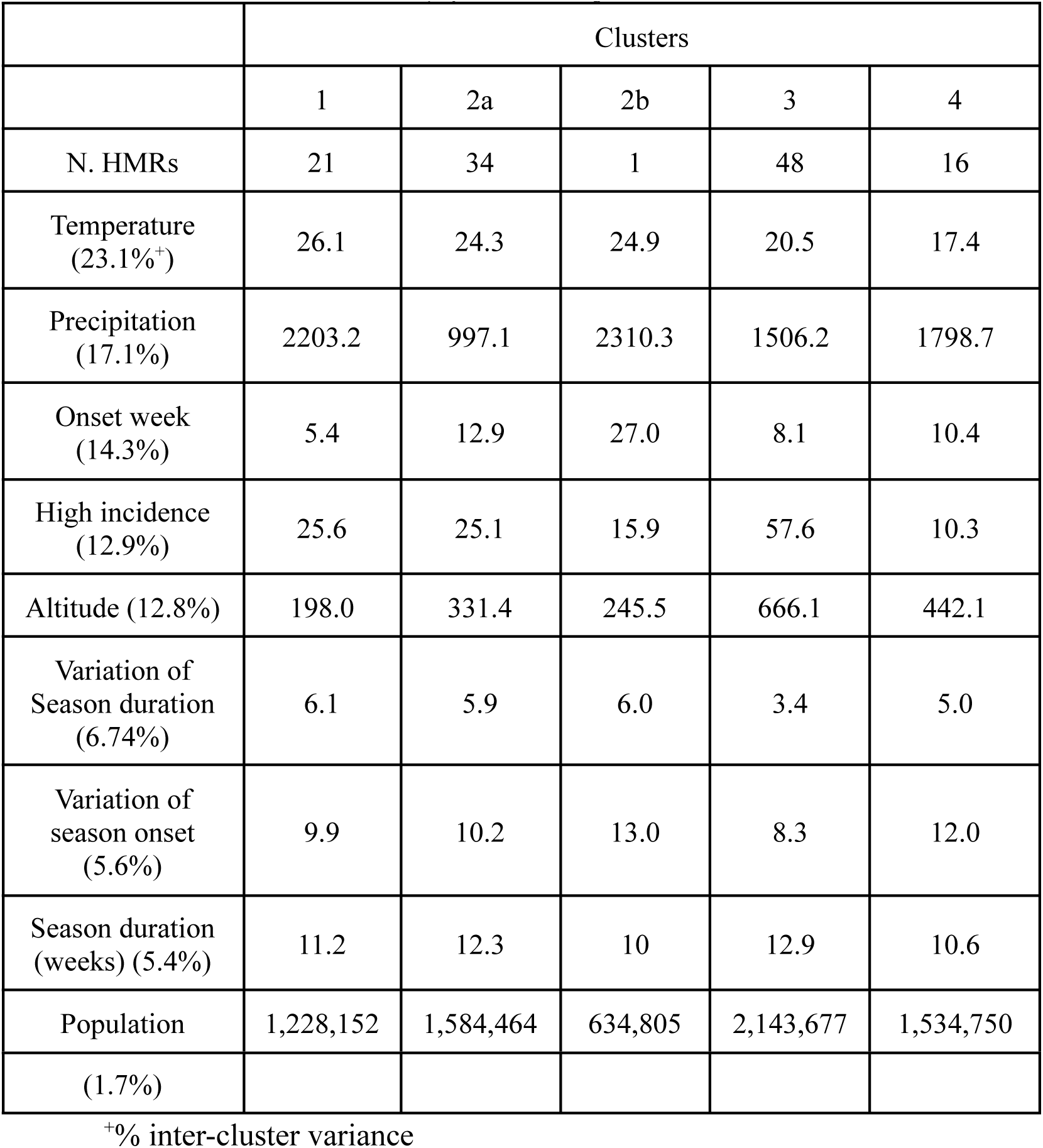
Mean values and relative contributions (%) of environmental and epidemiological variables to cluster differentiation, quantified by inter-cluster variance.

|  | Clusters |  |  |  |  |
| --- | --- | --- | --- | --- | --- |
|  | 1 | 2a | 2b | 3 | 4 |
| N. HMRs | 21 | 34 | 1 | 48 | 16 |
| Temperature (23.1%<sup>+</sup>) | 26.1 | 24.3 | 24.9 | 20.5 | 17.4 |
| Precipitation (17.1%) | 2203.2 | 997.1 | 2310.3 | 1506.2 | 1798.7 |
| Onset week (14.3%) | 5.4 | 12.9 | 27.0 | 8.1 | 10.4 |
| High incidence (12.9%) | 25.6 | 25.1 | 15.9 | 57.6 | 10.3 |
| Altitude (12.8%) | 198.0 | 331.4 | 245.5 | 666.1 | 442.1 |
| Variation of Season duration (6.74%) | 6.1 | 5.9 | 6.0 | 3.4 | 5.0 |
| Variation of season onset (5.6%) | 9.9 | 10.2 | 13.0 | 8.3 | 12.0 |
| Season duration (weeks) (5.4%) | 11.2 | 12.3 | 10 | 12.9 | 10.6 |
| Population | 1,228,152 | 1,584,464 | 634,805 | 2,143,677 | 1,534,750 |
| (1.7%) |  |  |  |  |  |

+% inter-cluster variance

The original data used for the Brazil climate classification was aggregated by HMR using the most frequent Köppen climate type as the predominant climatic classification (Table 2 in supplementary information 1). Additionally, from the same study, we used the available temperature, altitude, and precipitation data to calculate mean for each HMR (Figure 1C, D and E). The final map is shown in Figure 2.

**Figure 2.**
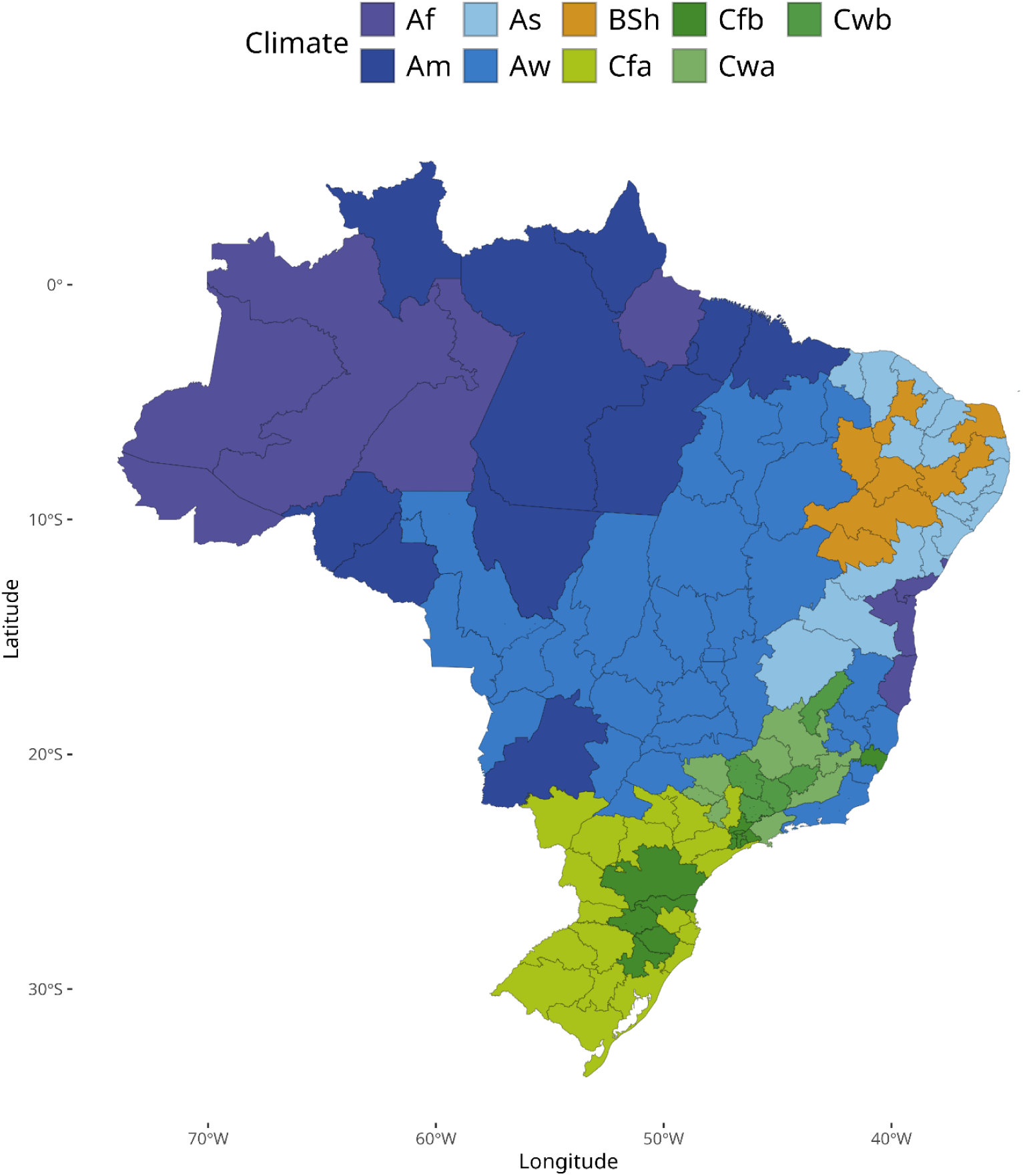
Köppen’s climate system of Brazil at HMR level derived from the original municipality classification by Alvares et al. (2013) (29). Service Layer Credits: Sources: https://www.ibge.gov.br/geociencias/organizacao-do-territorio/malhas-territoriais/15774-malhas.html?=&t=downloads. Köppen’s climate system of Brazil: Sources: https://www.ipef.br/publicacoes/acervohistorico/geodatabase/ as open access data and in https://zenodo.org/records/15281683 under a Creative Commons Attribution 4.0 International

### Moving Epidemics Method (MEM)

To extract the seasonal parameters for dengue, we applied the Moving Epidemics Method (MEM). This method estimates several parameters from the notification time series, including pre– and post-epidemic thresholds, typical activity levels, weekly expected incidence values, typical season onset, and duration (19,30). The method can be summarized into three main steps: First, the seasons are split into three periods: a pre-epidemic, an epidemic, and a post-epidemic period. The duration, onset, and end date of seasons are measured (Figure 3). Secondly, incidence thresholds are calculated using the pre– and post-epidemic incidences from historical seasons. Third, epidemic thresholds are computed within the epidemic season. The algorithm used is the one implemented in the R package “*MEM”* (31).

**Figure 3.**
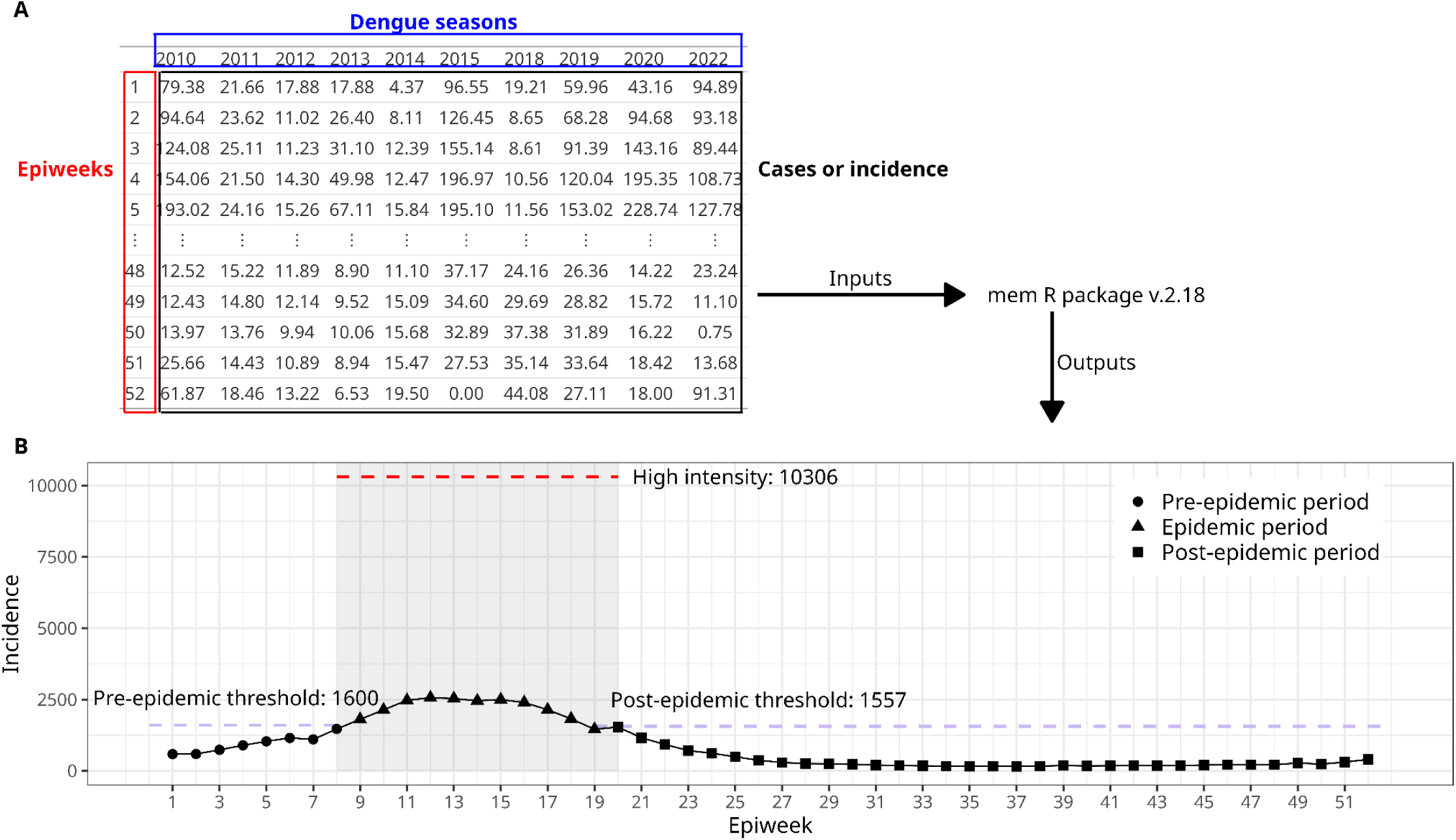
Diagram illustrating the MEM method applied to the HMR RRAS 12, in São Paulo state; A: The data input contains the number of cases per week per year, needed to describe the disease’s historical pattern (the dengue seasons used were those inside the percentiles 10% and 90%); B: MEM outputs: The year is split into three periods: pre-epidemic, epidemic and post-epidemic period, with their corresponding dates, duration and incidence thresholds. Intensity levels are computed based on historical quantiles. The grey region represents the epidemic period; red dashed line marks the high-intensity period.

Differently from the standard use of MEM, here, we introduced a pre-processing step. In the original MEM, extreme seasons are removed before analysis, manually. Here we implemented an algorithm for selecting these extreme seasons for exclusion. For each HMR, the geometric mean incidence of each season is calculated; then the geometric distance of each season’s peak to the geometric mean is computed. Atypically extreme seasons were identified as those exceeding the 10th and 90th percentiles of the entire geometric distance distribution (more information in supplementary material 2).

After this pre-processing, the MEM metrics are calculated as follows, for each HMR, using weekly dengue count data from 2010 to 2023:

#### Typical duration, onset, and end season dates

The length of the epidemic period is measured using the Maximum Accumulated Percentage approach (MAP), as described in (32,33). It defines the epidemic season as starting at week *k*, where MAP is maximized, and finishing at week *k + r – 1*, where *r* is the number of weeks used for maximization. Accordingly, the pre-epidemic period corresponds to the preceding weeks (1 to *k – 1*), while the post-epidemic period includes subsequent weeks (*k + r* to 52). Confidence intervals (95% CI) for duration, onset and end date were obtained from the standard bootstrap routine implemented in the MEM package. From that, we calculated the variation of these estimates, as the difference between their upper and lower bounds.

#### Pre-epidemic and pos-epidemic incidence thresholds

The pre-epidemic threshold is defined as the incidence above which an outbreak typically occurs in a specific region, where an outbreak is a sustained increase in cases greater than sporadic fluctuations. This threshold is estimated from the case distribution during the pre-epidemic period along all evaluated seasons and corresponds to the upper limit of the 95% confidence interval for this distribution. Incidence values below this threshold are assumed to be random fluctuations from a baseline. The post-epidemic threshold is calculated similarly, based on post-epidemic period data.

#### Intensity levels

This method also classifies the case volume per week as baseline, low, moderate, high, or very high compared to historical data (19). The baseline level falls below the pre-epidemic threshold; the low activity ranges from the epidemic threshold and continues until the 40% quantile. Moderate intensity ranges from 40% to 90%, and high activity from 90% to 97.5%. Values above 97.5% are classified as very high activity (Figure 3).

### Cluster analysis

Initially, a correlation analysis was conducted to address multicollinearity within the dataset. A cutoff of 0.8 for the Pearson correlation index revealed a strong correlation among the following variables: pre-epidemic threshold, post-epidemic threshold, and high intensity level threshold. Preliminary analyses comparing clustering results with each of these variables separately show best results with the high-intensity level threshold, and these are the results presented in the main text. The preliminary analyses using pre-epidemic or post-epidemic thresholds instead of the high-intensity level are shown in supplementary information 3.

Cluster analysis was performed using the following variables from MEM, after standardizing by the Z-Score method: high-intensity level, season onset date (mean and variation), and typical season duration (mean and variation). Additionally, the following environmental variables were included: mean annual temperature, mean annual precipitation, and mean altitude. The k-means clustering algorithm was chosen due to its widely used in scientific literature, computation efficiency, and straightforward implementation (34,35).

To determine the optimal number of clusters, both the elbow and silhouette methods were employed (36). The latter was also used to evaluate cluster quality, where a higher mean silhouette width indicated better-defined clusters. To help the characterization of the clusters, we computed the average of each variable, by cluster (37). Additionally, the variables that most contributed for cluster differentiation were identified, based on the between-centroid variance. The proportion of total variance explained by each variable was then computed by dividing the variable’s variance by the total variance of the dataset. This approach highlights the variables that play the most significant role in distinguishing the clusters. Finally, maps were produced to visualize the cluster classification for each region.

#### Post-processing

HMRs that showed significant discrepancies from their original cluster classification were manually reassigned to clusters based on MEM outputs, such as mean outbreak onset or high-intensity levels, that better aligned with their characteristics. When HMRs were not compatible with any existing clusters, a new cluster was created. This approach ensured that HMRs with distinct historical transmission patterns were not aggregated into the same cluster, thus avoiding the grouping of HMRs with high dengue occurrence alongside those with low or rare occurrence or distinct outbreak timing.

## Results

### Description of typical dengue seasons

From a total of 1,560 seasons (14 years x 120 HMRs), 1,246 (79.93%) were classified as typical. These are seasons whose geometric mean incidence fell within the 10th and 90th percentiles of the geometric distribution (0.27 – 3.45). The majority of HMRs presented at least five typical seasons, with 66% of them with 10 to 13 seasons, while only 8.46% with less than 8. There were two HMRs in the subtropical region (North and Northeast Santa Catarina state) that recorded only three seasons. To address this limitation, we included seasons with geometric distances closer to the predefined cutoff percentiles, to reach the minimum of 5 seasons.

#### Pre-epidemic and post-epidemic thresholds

The overall mean pre-epidemic threshold was 3.26 cases per 100,000 inhab/week (95% CI: 2.66 – 4.01) while the post-epidemic threshold was 5.62 cases per 100,000 inhab/week (95% CI: 4.76 – 6.48). The highest values of both metrics were observed in the westernmost HMR (Acre), and in the Central-West, where some HMRs exhibited incidence thresholds up to three times higher than the overall mean. Examples are the Central-West HMR in Góias state and RRAS 12 in São Paulo state. In contrast, low incidence thresholds were found in the South and Northeast regions (Figure 4A, B). Low post-epidemic threshold values are observed in the Southern HMRs.

**Figure 4.**
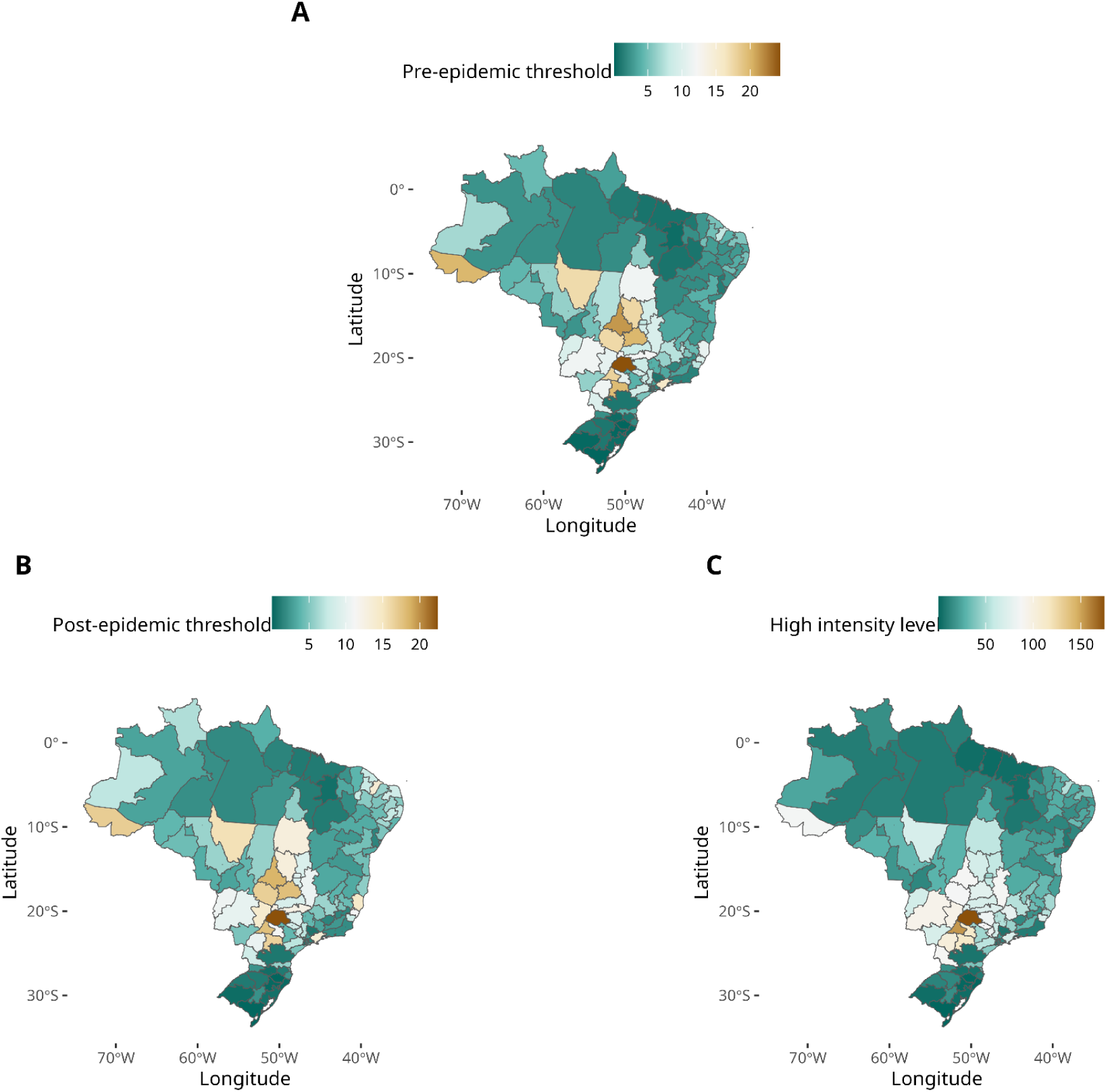
Characteristics of typical dengue seasons in the Brazilian health macroregions; A: pre-epidemic threshold is defined as the incidence that marks the beginning of the season (cases/week x 10^5); B: post-epidemic threshold is the incidence that marks the end of the season (cases/week x 10^5); C: High-intensity level is the incidence threshold above which a season is considered a severe epidemic (cases/week x 10^5). Service Layer Credits: Sources: https://www.ibge.gov.br/geociencias/organizacao-do-territorio/malhas-territoriais/15774-malhas.html?=&t=downloads

#### High-intensity threshold level

The high-intensity threshold level averaged 36.22 cases per 100,000 inhab/week (95% CI: 30.32 – 42.12) among the HMRs. The highest values (> 150) are seen in São Paulo and Paraná states, in the Southeast region. Moderate values (between 50 and 150) are found in Acre and in the Center-West region, while lower levels (< 50) were found throughout the country, especially in the Southern states, such as Rio Grande do Sul and Santa Catarina (Figure 4C) and in some areas of the North.

#### Season onset and duration

The mean start date of the dengue seasons in Brazil is epiweek 9 (95% CI: 8 – 10) but there is a high regional variability (Figure 5A). The earlier onsets occur in the west (longitude 60-70 W), up to week 5. Meanwhile, the eastern HMRs (longitude 40 – 45 W) present later onsets, at week 15 to 25. A deviation from this west-east pattern is Roraima (the northernmost HMR) which shows a late onset at week 25. This is the only HMR located in the Northern hemisphere. At mid-latitudes, there is more variability, with a few HMRs in the Northwest and Central-South regions (12.5%) tending to experience earlier onsets, while late start is observed in some HMRs in the South and Southeast regions. (Figure 5A).

**Figure 5.**
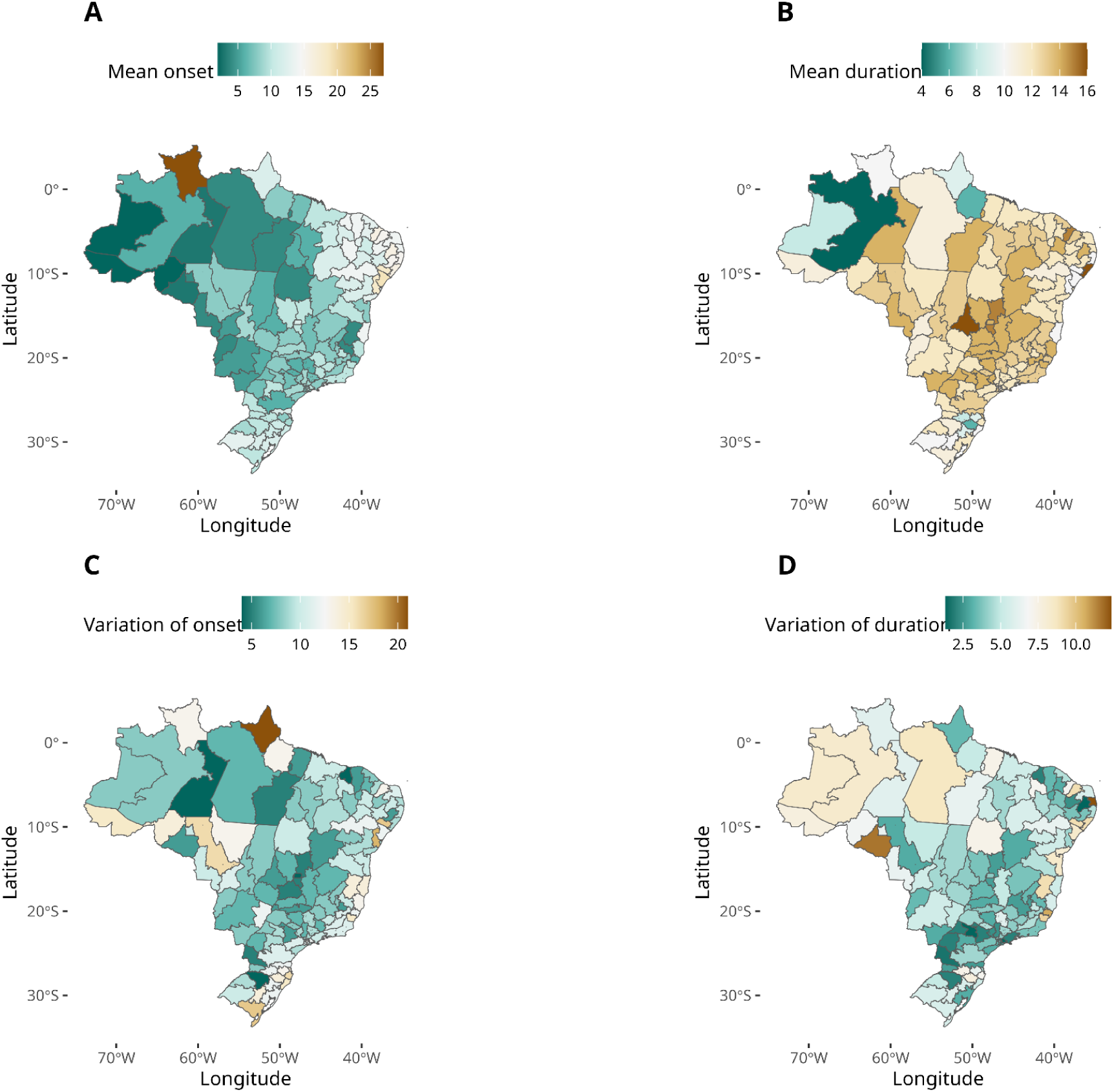
Characteristics of typical dengue seasons in the Brazilian health macroregions; A: Season onset date is the epidemiological week at which the pre-epidemic threshold is surpassed.; B: Mean season duration is the number of weeks with incidence above the pre-epidemic threshold; C: variation of onset date, measured as the width of the onset’s 95% confidence interval; D: variation of the season duration, measured as the width of the season duration 95% confidence interval. Service Layer Credits: Sources: https://www.ibge.gov.br/geociencias/organizacao-do-territorio/malhas-territoriais/15774-malhas.html?=&t=downloads

The variation of the estimated onset date reflects the persistence of the seasonal period. We observed lower onset variation in the North and Central-West HMRs, suggesting a marked seasonality. In contrast, there are HMRs showing variation exceeding 15 epiweeks, particularly in the Southeast, South, and North regions, with the HMR of Amapá state exhibiting the greatest uncertainty (Figure 5B).

The mean duration of a dengue season in Brazil is 12 weeks (95% CI: 11 – 13). Among all HMRs, 93.3% exhibited seasons exceeding 10 weeks. Only the central and western HMRs in the Amazonas state, the Amapá HMR, and HMR I in Pará presented seasons less than 10 weeks long (Figure 5C). These are all located in the tropical north. Concerning the variation of the estimated season durations, 56.6% of all HMRs exhibited variation less than 5 weeks, indicating a high inter-year consistency. The highest uncertainty was observed in the HMRs of the Northeast and North regions (Figure 5D).

#### Cluster analysis

The optimal number of clusters (k), as determined by the elbow method, was assessed to be 3 or 4. A silhouette plot with k = 4 demonstrated a higher average silhouette width and less overlap between clusters compared to other configurations (Figure 6), thereby justifying its selection as the preferred model. Additional results utilizing k = 3 to 6 are provided in supplementary Information 2. Overall, increasing k resulted in a decrease in the mean silhouette width, suggesting the possibility of misclassification (Figure 6C and D). The variables most effective in distinguishing between clusters, as indicated by the variation among cluster centroids, were mean temperature, mean precipitation, mean onset week, high-intensity incidence threshold, and mean altitude. These variables exhibited the greatest variance among clusters. Collectively, they account for 80% of the total variance between the cluster centroids.

**Figure 6.**
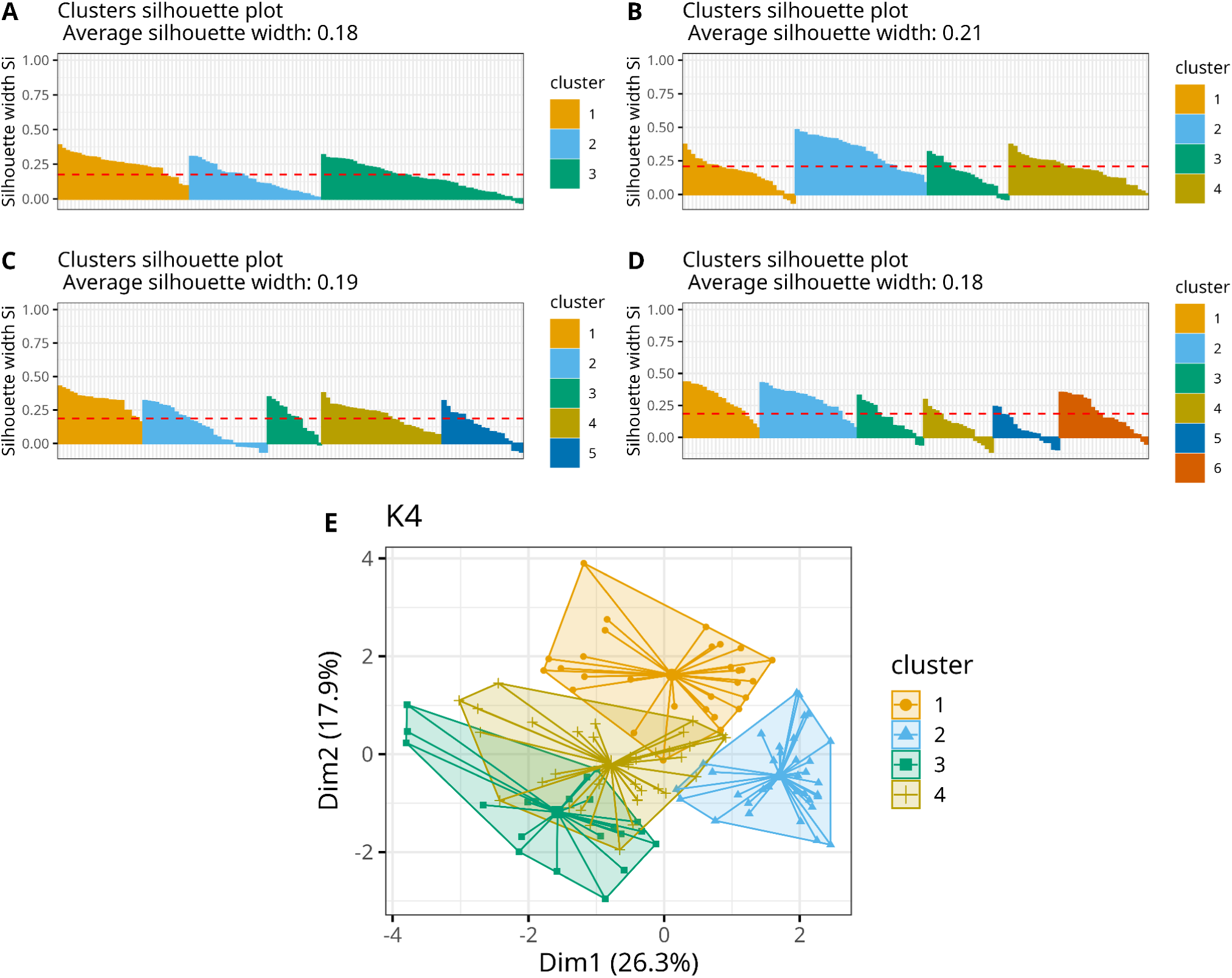
Results of the multivariate cluster analysis for the classification of health macroregions into homogeneous groups based on their climate and dengue transmission seasons; (A-D) silhouette plot for analysis considering 3 to 6 clusters. (E) Cluster plot using 4 clusters, the final classification.

Figure 7A illustrates the spatial distribution of the four clusters. HMRs belonging to the same clusters are also spatially clustered, reflecting similarities in their climate characteristics. Cluster 1 encompasses approximately 50% of the country, composed primarily by HMRs in the North, and a few located in the Center-West. Cluster 2 covers a significant area in the east, as well as a non-contiguous HMR in the north (Roraima). Given the substantial geographical separation and documented socio-environmental differences between these areas, we further subdivided Cluster 2 into two distinct subclusters (2a and 2b). Cluster 3 is situated in the central part of the country, extending further south compared to the previous clusters. Finally, Cluster 4 comprises the southern HMRs and a non-contiguous area in the southeast. These clusters delineate regions with distinct characteristics related to climate, as well as the timing and intensity of dengue outbreaks.

**Figure 7.**
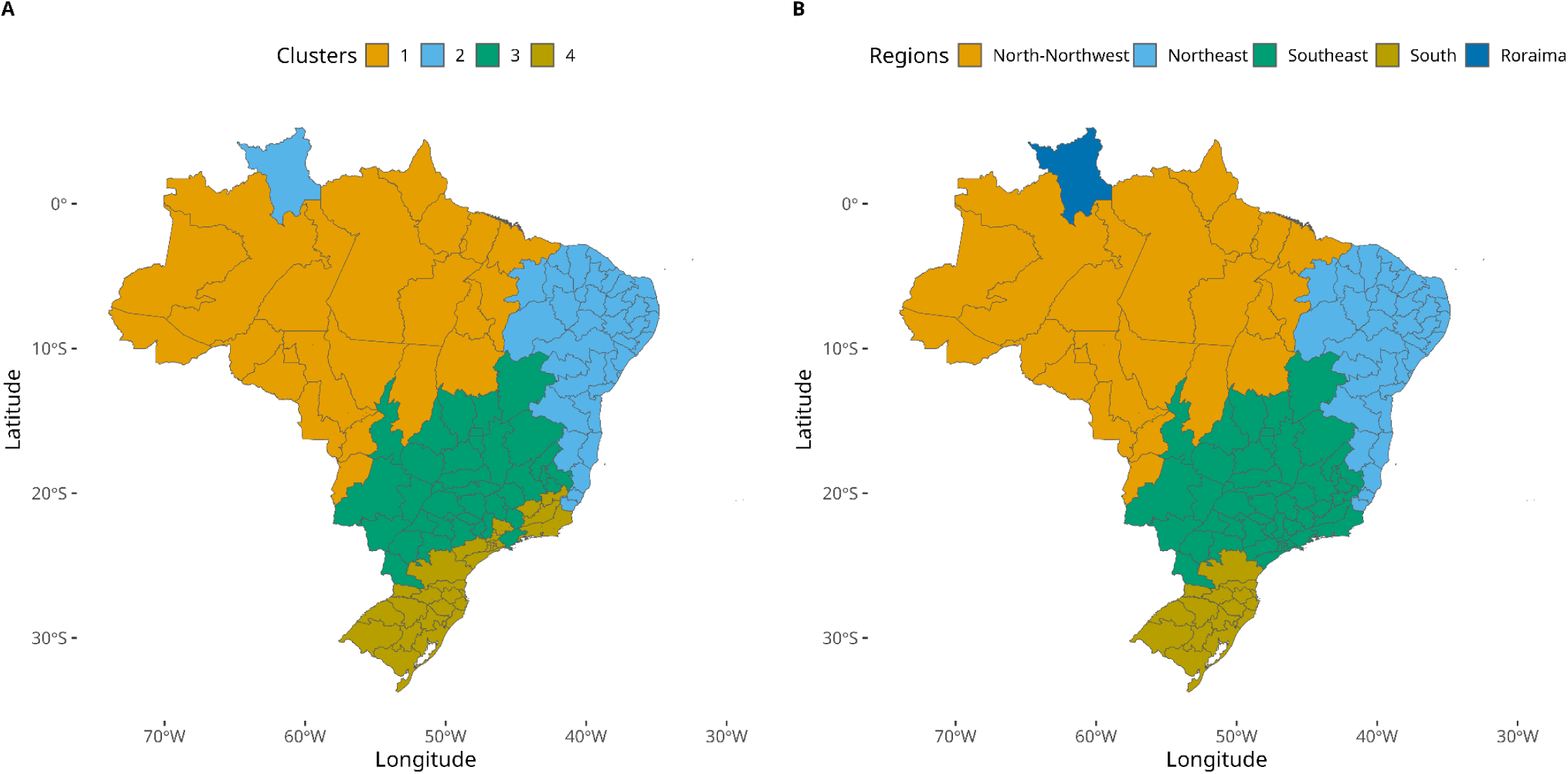
Health Macro Region Clustering and Final Dengue Regionalization of Brazil; (A) Classification of HMR into 4 clusters, following the k-means algorithm; (B) Final classification of Brazil into 5 dengue regions, after the post processing phase. Service Layer Credits: Sources: https://www.ibge.gov.br/geociencias/organizacao-do-territorio/malhas-territoriais/15774-malhas.html?=&t=downloads

Temperature was the primary determinant variable for cluster differentiation, explaining 23.1% of the inter-cluster variance (Table 1). This thermal influence is evident in the warm-to-mild gradient observed across clusters 1 through 4, following distinct latitudinal and longitudinal patterns. Precipitation poses as the second strongest differentiation variable (17.1% of variance), with values decreasing from clusters 1 and 2b (highest) through clusters 3 and 4 (intermediate) to cluster 2a (lowest), consistent with the latter’s semi-arid characteristics.

Among MEM-derived metrics, seasonal onset week was the most important for differentiation (14.3% of variance), revealing a clear spatiotemporal progression: epidemic timing began earliest in tropical cluster 1 (late January to early February), followed sequentially by subtropical cluster 3 (mid-February), temperate cluster 4 (early March), semi-arid cluster 2a (mid-March), and finally highland cluster 2b (June). The high-intensity incidence threshold contributed substantially (12.9%), while seasonal duration and onset variability showed more modest effects (6.7% and 5.6% of variance, respectively).

### Dengue regionalization in Brazil

The proposed regionalization of typical dengue seasons is based on the results of the cluster analysis, with a few adjustments, to avoid discontinuous regions. First, we implemented the above mentioned split of cluster 2 into two. Second, the small cluster of 14 HMRs in the Southeast, initially classified as cluster 4 (in Rio de Janeiro, Minas Gerais, and São Paulo states) were reassigned to cluster 3. This choice is justified by the closer administrative relationship between these HMRs and their neighbors in cluster 3. As a result, cluster 4 now comprises only the HMRs in the subtropical South region, characterized by colder temperatures and historically lower numbers of cases compared to other HMRs. The final classification map is shown in Figure 7B.

## Discussion

The classification of the country into five regions, based on climatic and dengue epidemiological data, provides a foundation for regionalized decision-making aimed at reducing the occurrence and impact of dengue outbreaks. Region 1 (North-Northwest), predominantly tropical, experiences outbreaks earlier than other regions. This may be attributed to the precipitation volume and year-around warm temperatures, both conducive to *Aedes* mosquito reproduction and survival (8,38). The effect of lower precipitation on outbreak onset is evident in Region 2a (NorthEast), which consists of HMRs with dry climates and tropical climates with dry summers, leading to later outbreak occurrences. Chronic water scarcity in this region encourages water storage, which, if not properly managed, can create favourable conditions for mosquito breeding (12), and potentially sustaining dengue transmission throughout the year (37). The most delayed season onset is observed in Region 2b (Roraima), composed by a single HMR. The distinct disease pattern in this region has been described elsewhere (17) and may be influenced by the local monsoon season that occurs in the later months. High incidence threshold, a defining characteristic of Region 3 (Southeast), reflects the high population density observed in the most populated cities of the country (39). Region 4 (South) has witnessed, in recent years, an increased transmission, with high incidence rates possibly due to a large portion of the population being naive to these arboviruses (40). This subtropical climate region has historically exhibited temperature ranges that limit mosquito population densities, particularly during winter months, resulting in consistently low dengue case reports. However, recent years have seen a marked expansion of dengue transmission into higher latitudes—a shift likely driven by climate change impacts on vector ecology and viral transmission dynamics. (41,42).

Various studies have investigated dengue transmission patterns, using methodologies such as time series decomposition, lag non-linear models, and spatial modeling to investigate the effects of outbreak frequencies, disease-free periods, hydrometeorological events, socio-environmental factors on transmission, prior outbreaks, population density, human mobility, number of months with temperatures suitable for *Aedes* mosquito activity (17,37,41,43–47). However, this study is the first to apply MEM in the context of a mosquito-transmitted viral disease, specifically to distinguish regions based on the combination of its results and environmental data.

The classification presented here can act as a reference for determining the optimal timing of vector control interventions across different regions. Given that outbreak onset typically varies between regions, the timing of such interventions should be region-specific. Despite uncertainties surrounding the precise onset of the epidemic week, these guidelines can help mitigate outbreaks by reducing mosquito populations to levels insufficient to sustain transmission.

Accordingly, control efforts should be prioritised to be initiated in Regions North-Northwest and West, followed by implementation in Regions Southeast and South, and, at last in Northeast and Roraima. These recommendations are particularly valuable given that newly implemented vector control strategies, including mosquito-disseminated pyriproxyfen (MDPPF) (48), do not produce an immediate reduction in adult mosquito populations (49), and the eco-health approach, which involves householders and the community as active participants in mosquito control, could also benefit from more accurate outbreak onset timing to inspect unprotected water storage, garbage, and potential breeding sites. In general, all aspects of managing mosquito-transmitted viral diseases can be improved by estimating the outbreak onset, including acquiring resources such as larvicides, providing educational materials to the community, and preparing health units for a potential rise in cases.

Similar to other methodologies that depend on notification data, the MEM results are subject to limitations, including delays in case reporting and the potential case misdiagnosis as other mosquito-borne viral diseases (49). Moreover, the approach is based on typical seasons, and the presence of new serotypes or climate variability, such as El Nino effects, can anticipate these seasons. In this case, continuous monitoring is necessary to adjust the expectations. Infodengue and Early Warning, Alert and Response System (EWARS) are examples of such systems.

While the Köppen climate classification remains a robust and widely used methodology, climate change continues to be a significant issue, contributing to factors such as pathogen spillover, increased transmission of waterborne diseases, and the expansion of areas suitable for vector distribution, thus increasing the risk of disease transmission in previously unaffected regions (50–54). The climate data used to characterize Brazil (1950–1990) may underestimate the effects of climate change on dengue transmission, as recent studies have shown the impact of climate anomalies on transmission, and their increasing frequency globally (40,55,56). These observations align with global warming trends, as the planet’s 10 warmest years on record have occurred in the last decade. Notably, 2024 marked the latest record, with an average global temperature 1.5°C higher than the 20th-century average, closely followed by 2023 (56). These rising temperatures are closely associated with an increase in the frequency and intensity of climate anomalies, such as extreme heatwaves, erratic rainfall patterns, and prolonged droughts (57,58). Together, these trends underscore climate change’s expanding influence on dengue transmission dynamics and the urgent need for adaptive public health strategies..

Classifying the HMRs into similar regions using epidemiological and climate data, offers a valuable framework for decision-makers to estimate the optimal timing for implementing control strategies targeting the *Aedes* mosquito vector control. While the Köppen climate classification system does not account for the recent effects of climate change, and disease data have inherent limitations of the notification delay, this approach provides a practical starting point for identifying regional patterns. However, some HMRs may share characteristics with multiple clusters and cannot be entirely classified into a single cluster using algorithms.

Despite these limitations, applying the mitigation and control efforts of mosquito-transmitted viral diseases at the timing indicated in this study can be beneficial, even in the face of uncertainty regarding outbreak onset, by reducing mosquito densities and potentially mitigating unforeseen outbreaks. This approach is particularly relevant given recent evidence demonstrating persistent outbreak risks following the unprecedented case numbers observed in 2024 (59). The proposed methodology can also be applied to other mosquito-borne viral diseases, including Zika and chikungunya, enhancing its public health impact. Future studies should investigate the classification process at a finer scale, even when focused on a specific region of the country, where the number of cases allows for a more detailed local-scale analysis. For this, the MEM method needs adaptation to work with small numbers of cases. Furthermore, applying the MEM to other arboviruses and comparing results with alternative regionalization approaches could enhance our understanding of outbreak patterns across different disease systems.

## Data Availability

All data produced in the present study are available upon reasonable request to the authors

## Acknowledgements

We thank the Infodengue team for the technical and scientific discussions on mosquito-transmitted virus disease surveillance in Brazil.

## Support Information Caption

**S1 Table 1.** Summary of climate types and subtypes per municipalities

**S1 Table 2.** Summary of climate types and subtypes per Health Macro-Regions (HMRs)

**S2 Figure 1**. Number of HMRs with different quantities of seasons inside the percentiles 10 and 90. A: After filtering, 1 HMR – Planalto Norte and Nordeste in the state of Santa Catarina – had lower than 5 seasons; B: Inclusion of seasons with geometric distance closest to the 10th and 90th percentiles.

**S3 Figure 1**. Correlation analysis with the full dataset. Variables with Pearson correlation index greater than equal –0.8 or 0.8 were removed to mitigate distortions in t Euclidean distance calculations on further cluster analysis. Thus, only one variable from the set of pre-epidemic threshold, post-epidemic threshold, and high-intensity level was kept for further analysis.

**S3 Figure 2**. The optimal number of clusters minimizes the within-cluster sum of squares by the elbow (large plots) and silhouette (small plots) methods. A) Using pre-epidemic threshold; B) Using post-epidemic threshold; C) Using high-intensity level. For both the pre-epidemic and post-epidemic thresholds, the silhouette method suggests 3 clusters, while 4 clusters are indicated for the high-intensity level.

**S3 Figure 3**. Cluster plots using the pre-epidemic threshold. The k-means algorithm found similarities between several HMRs but it’s possible to note a lot of overlap between the clusters. A-D: 3-6 clusters.

**S3 Figure 4**. Cluster quality using the pre-epidemic threshold and different numbers of clusters. A-D: 3-6 clusters. The increase in the number of clusters decreases the mean silhouette indicating possible misclassifications.

**S3 Figure 5**. Maps generated with the cluster analysis with a pre-epidemic threshold. A-D: 3-6 Clusters. Service Layer Credits: Sources: https://www.ibge.gov.br/geociencias/organizacao-do-territorio/malhas-territoriais/15774-malhas.html?=&t=downloads

**S3 Figure 6**. Cluster plots using the post-epidemic threshold. The k-means algorithm found similarities between several HMRs but it’s possible to note a lot of overlap between the clusters. A-D: 3-6 clusters.

**S3 Figure 7**. Cluster quality using the post-epidemic threshold and different numbers of clusters. A-D: 3-6 clusters. The increase in the number of clusters decreases the average silhouette indicating possible misclassifications.

**S3 Figure 8**. Maps generated with the cluster analysis with a pre-epidemic threshold. A-D: 3-6 Clusters. Service Layer Credits: Sources: https://www.ibge.gov.br/geociencias/organizacao-do-territorio/malhas-territoriais/15774-malhas.html?=&t=downloads

**S3 Figure 9**. Cluster plots using the high-intensity level. The k-means algorithm found similarities between several HMRs but it’s possible to note a lot of overlap between the clusters. A-C: 3, 5 and 6 clusters. The plot with 4 clusters is in the main document.

**S4 – Figure 1.** Number of dengue cases per 100,000 inhabitants for each Health Macro-Regions (HMR) in 2010. Service Layer Credits: Sources: https://www.ibge.gov.br/geociencias/organizacao-do-territorio/malhas-territoriais/15774-malhas.html?=&t=downloads

**S4 – Figure 2** Number of dengue cases per 100,000 inhabitants for each Health Macro-Regions (HMR) in 2011. Service Layer Credits: Sources: https://www.ibge.gov.br/geociencias/organizacao-do-territorio/malhas-territoriais/15774-malhas.html?=&t=downloads

**S4 – Figure 3** Number of dengue cases per 100,000 inhabitants for each Health Macro-Regions (HMR) in 2012. Service Layer Credits: Sources: https://www.ibge.gov.br/geociencias/organizacao-do-territorio/malhas-territoriais/15774-malhas.html?=&t=downloads

**S4 – Figure 4** Number of dengue cases per 100,000 inhabitants for each Health Macro-Regions (HMR) in 2013. Service Layer Credits: Sources: https://www.ibge.gov.br/geociencias/organizacao-do-territorio/malhas-territoriais/15774-malhas.html?=&t=downloads

**S4 – Figure 5** Number of dengue cases per 100,000 inhabitants for each Health Macro-Regions (HMR) in 2014. Service Layer Credits: Sources: https://www.ibge.gov.br/geociencias/organizacao-do-territorio/malhas-territoriais/15774-malhas.html?=&t=downloads

**S4 – Figure 6** Number of dengue cases per 100,000 inhabitants for each Health Macro-Regions (HMR) in 2015. Service Layer Credits: Sources: https://www.ibge.gov.br/geociencias/organizacao-do-territorio/malhas-territoriais/15774-malhas.html?=&t=downloads

**S4 – Figure 7** Number of dengue cases per 100,000 inhabitants for each Health Macro-Regions (HMR) in 2016. Service Layer Credits: Sources: https://www.ibge.gov.br/geociencias/organizacao-do-territorio/malhas-territoriais/15774-malhas.html?=&t=downloads

**S4 – Figure 8** Number of dengue cases per 100,000 inhabitants for each Health Macro-Regions (HMR) in 2017. Service Layer Credits: Sources: https://www.ibge.gov.br/geociencias/organizacao-do-territorio/malhas-territoriais/15774-malhas.html?=&t=downloads

**S4 – Figure 9** Number of dengue cases per 100,000 inhabitants for each Health Macro-Regions (HMR) in 2018. Service Layer Credits: Sources: https://www.ibge.gov.br/geociencias/organizacao-do-territorio/malhas-territoriais/15774-malhas.html?=&t=downloads

**S4 – Figure 10** Number of dengue cases per 100,000 inhabitants for each Health Macro-Regions (HMR) in 2019. Service Layer Credits: Sources: https://www.ibge.gov.br/geociencias/organizacao-do-territorio/malhas-territoriais/15774-malhas.html?=&t=downloads

**S4 – Figure 11** Number of dengue cases per 100,000 inhabitants for each Health Macro-Regions (HMR) in 2020. Service Layer Credits: Sources: https://www.ibge.gov.br/geociencias/organizacao-do-territorio/malhas-territoriais/15774-malhas.html?=&t=downloads

**S4 – Figure 12** Number of dengue cases per 100,000 inhabitants for each Health Macro-Regions (HMR) in 2021. Service Layer Credits: Sources: https://www.ibge.gov.br/geociencias/organizacao-do-territorio/malhas-territoriais/15774-malhas.html?=&t=downloads

**S4 – Figure 13** Number of dengue cases per 100,000 inhabitants for each Health Macro-Regions (HMR) in 2022. Service Layer Credits: Sources: https://www.ibge.gov.br/geociencias/organizacao-do-territorio/malhas-territoriais/15774-malhas.html?=&t=downloads

**S4 – Figure 14** Number of dengue cases per 100,000 inhabitants for each Health Macro-Regions (HMR) in 2023. Service Layer Credits: Sources: https://www.ibge.gov.br/geociencias/organizacao-do-territorio/malhas-territoriais/15774-malhas.html?=&t=downloads

**S5** – Dataset used for multivariate cluster analysis

## Availability of data and material

The datasets generated and/or analysed during the current study are available in the supplementary material.

## Authors’ contributions

ASG, MFCG, IFA, RML, LSB, SSO, ECA, DACF, VBG, LBV, TISR, OGC, FCC, CTC, contributed to the main idea and structure of the project, the proposal and implementation of the MEM for dengue data, interpretation of the final data, and writing of the manuscript. ASG prepared the data and performed descriptive and multivariate analyses. All authors read and approved the final manuscript.

