## Supplementary information 1 for "Unraveling Regional Variability in Dengue Outbreaks in Brazil: leveraging the Moving Epidemics Method (MEM) and Climate Data to Optimize Vector Control Strategies"

Köppen Climate System of Brazil

**S1-Table 1.** Summary of climate types and subtypes per municipalities

| Abbreviation | Climate type | Subtype | Number of municipalities |
| --- | --- | --- | --- |
| Af | Tropical | Without dry season | 219 |
| Am |  | Monsoon | 397 |
| As |  | Dry winter | 891 |
| Aw |  | Dry summer | 1336 |
| Bsh | Dry | Semi-arid low altitude and latitude | 420 |
| Cfa | Humid subtropical | Hot summer | 1137 |
| Cfb |  | Temperate summer | 405 |
| Cwa |  | Dry winter and hot summer | 407 |
| Cwb |  | Dry winter and temperate summer | 355 |

**S1-Table 2.** Summary of climate types and subtypes per Health macro-region (HMR)

| Abbreviation | Climate type | Subtype | Number of HMR |
| --- | --- | --- | --- |
| Af | Tropical | Without dry season | 8 |
| Am |  | Monsoon | 10 |
| As |  | Dry winter | 17 |
| Aw |  | Dry summer | 31 |
| Bsh | Dry | Semi-arid low altitude and latitude | 8 |
| Cfa | Humid subtropical | Hot summer | 20 |
| Cfb |  | Temperate summer | 13 |
| Cwa |  | Dry winter and hot summer | 8 |
| Cwb |  | Dry winter and temperate summer | 5 |
