## Supplementary information 2 for "Unraveling Regional Variability in Dengue Outbreaks in Brazil: leveraging the Moving Epidemics Method (MEM) and Climate Data to Optimize Vector Control Strategies"

Filtering atypical dengue seasons

To increase the accuracy of MEM outputs, a prior dataset of municipality dengue incidence peaks of each season (**xp_df**) was aggregated to the Health Macro-Region (HMR) scale (**HMR_xp**) and used to guide the identification and filtering of atypical dengue seasons in the historical data of each HMR. In this way, the characterization of dengue seasons within each HMR as typical or atypical was based on the geometric distance of each season. This distance is calculated as the ratio of the incidence peak (**xp_macro**) for each season to the geometric mean of incidence peaks (**geom_mean**) across all seasons. Typical seasons are identified as those with geometric distances between the 10th (0.27) and 90th (3.45) percentiles of the overall distribution, while atypical seasons fall outside this range. The following R code chunk demonstrates this process.

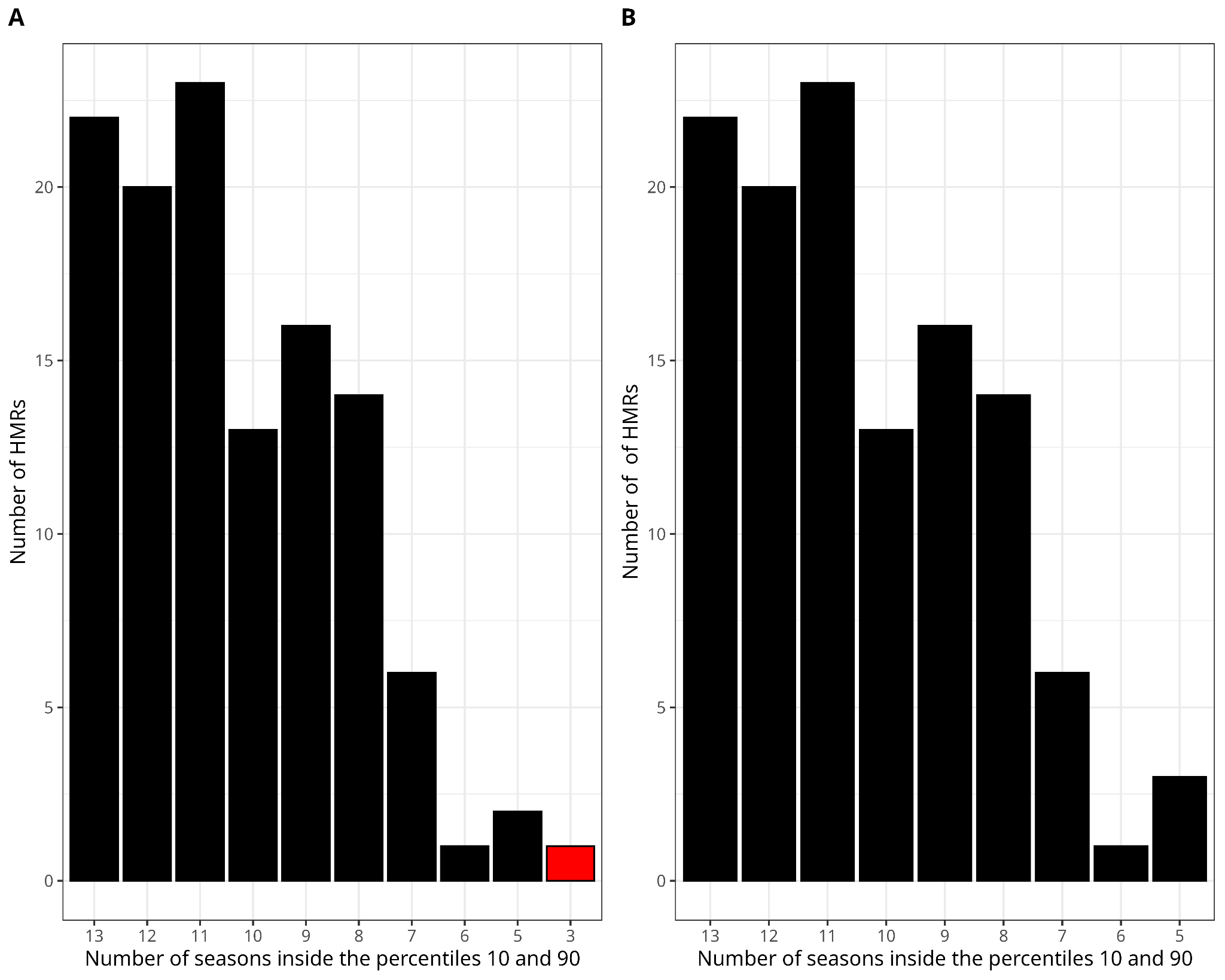

**S2-Figure 1** Number of HMRs with different quantities of seasons inside the percentiles 10 and 90. A: After filtering, 1 HMR - Planalto Norte and Nordeste in Santa Catarina state - had lower than 5 seasons; B: Inclusion of seasons with geometric distance closest to the 10th and 90th percentiles.

```
library(tidyverse)


mean_geom <- function(x) {
 exp(mean(log(x)))
}

# HMR data
HMR_df <- xp_df %>% left_join(macro_mun) %>%
           group_by(code_macro,ano_e,state) %>%
           summarise(xp_macro = sum(xp))

# calculating geom_mean and geom_distance
HMR_df <- HMR_df %>% filter(xp_macro!=0) %>%
           group_by(code_macro) %>%
           mutate(geom_mean = mean_geom(xp_macro),
             geom_distance = xp_macro / geom_mean)

percentiles10_90 <- quantile(dadosMC$distancia_geom,
                     probs=c(0.10, 0.90))

#only seasons in the percentiles
HMR_df2 <- HMR_df %>% group_by(code_macro) %>%
           filter(geom_distance >= percentiles10_90[1] &
             distancia_geom <= percentiles10_90[2])

#finding HMR without 5 seasons after filtering


HMR_is <- HMR_df2 %>% group_by(code_macro) %>% # HMR with insufficient seassons
  tally() %>%
  filter(n < 5) %>% mutate(n = 5-n)
```

To run the MEM model in each HMR, a minimum of 5 seasons was required to ensure a reasonable amount of data. After filtering, 51.16% of HMRs had more than 10 typical seasons included (Figure 1A). Only one HMR—Planalto Norte and Nordeste in the state of Santa Catarina—had fewer than 5 seasons remaining after the filtering process, with only the seasons of 2014, 2015, and 2018 originally falling within the 10th and 90th percentiles. To address this issue, for HMRs with fewer than 5 seasons after filtering out atypical ones, additional seasons with geometric distances closest to the 10th and 90th percentiles were included (Figure 1B). The R code chunk below illustrates this process.

```
#iterate to each HMR with insufficient number of seasons
for (i in seq_len(nrow(HMR_is))) {
  code_HMR_miss <- HMR_is$code_macro[i]
  necessary_seasons <- HMR_is$n[i]

#selecting seasons with geom_distance near the percentiles
 HMR_df3 %>% filter(code_macro == code_HMR_miss,
              geom_distance < percentiles10_90[1] |
              geom_distance > percentiles10_90[2]) %>%
             arrange(code_macro, abs(distancia_geom - percentiles10_90[1]),
              abs(geom_distance -percentiles10_90[2])) %>%
             slice_head(n = necessary_seasons)

}

#data with at least 5 seasons to all HMRs used in mem model
HMR_ok <- bind_rows(HMR_df2, HMR_df3)
```

This filtering process of atypical seasons is currently used in the routine of Brazilian surveillance of severe acute respiratory syndrome (SARS) and influenza through the Infogripe system (<https://info.gripe.fiocruz.br>), as well as for dengue, Zika, and chikungunya surveillance through the Infodengue system (<https://info.dengue.mat.br>).
