## Supplementary information 3 for "Unraveling Regional Variability in Dengue Outbreaks in Brazil: leveraging the Moving Epidemics Method (MEM) and Climate Data to Optimize Vector Control Strategies"

Cluster analysis complete results

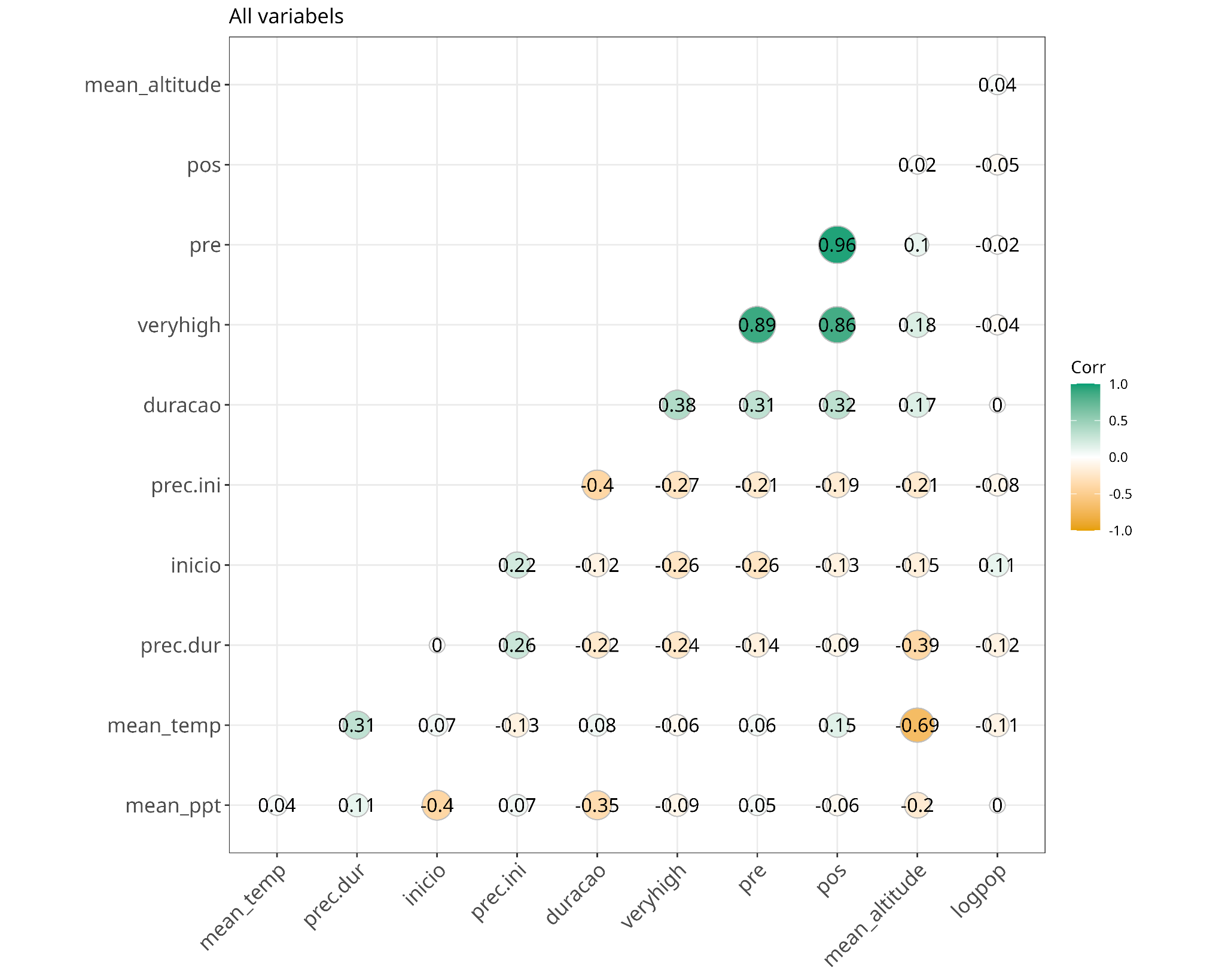

**S3-Figure 1**. Correlation analysis with the full dataset. Variables with Pearson correlation index greater than equal -0.8 or 0.8 were removed to mitigate distortions in t Euclidean distance calculations on further cluster analysis. Thus, only one variable from the set of pre-epidemic threshold, post-epidemic threshold and high-intensity level was kept for further analysis.

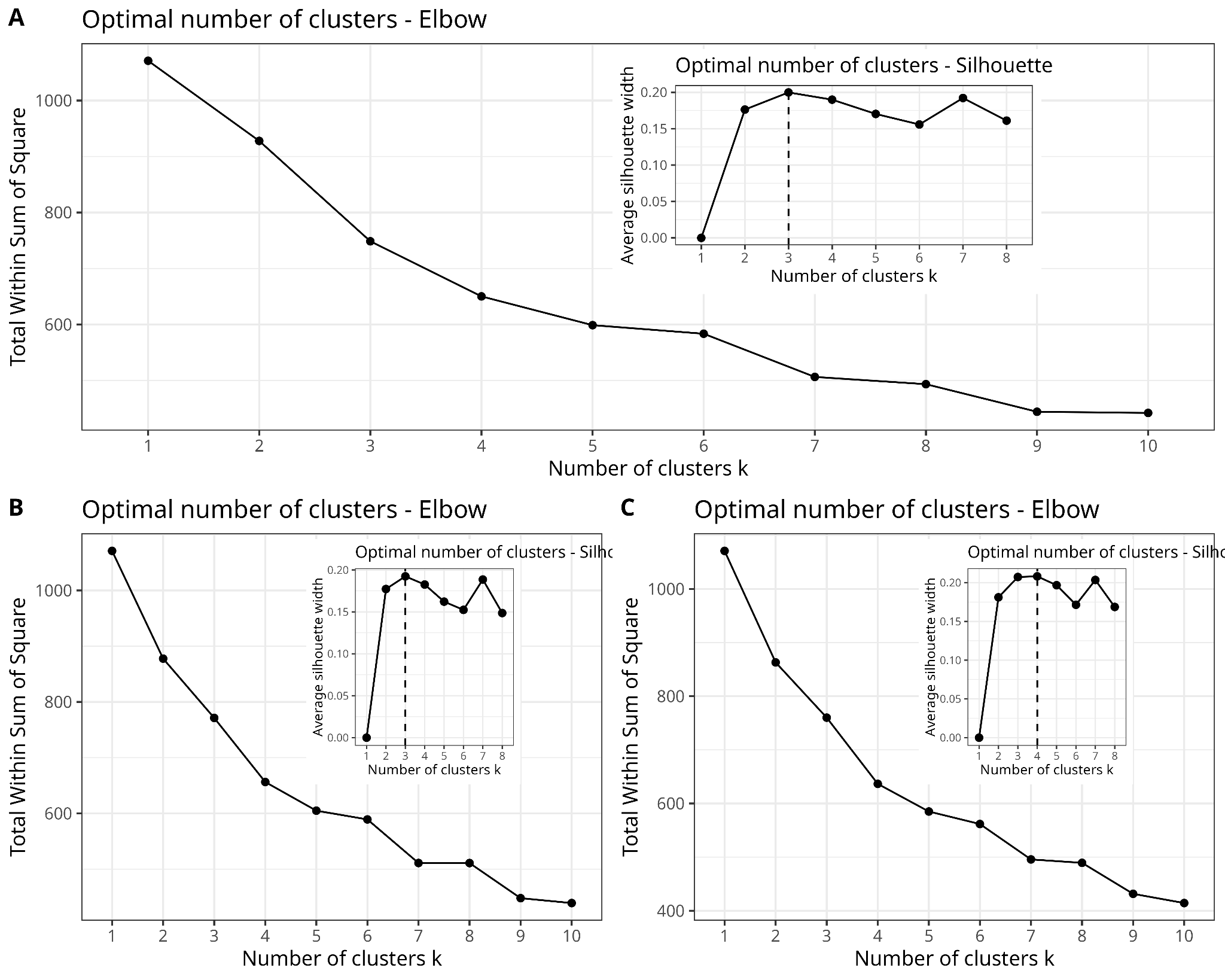

**S3-Figure 2**. The optimal number of clusters minimizes the within-cluster sum of squares by the elbow (large plots) and silhouette (small plots) methods. A) Using pre-epidemic threshold; B) Using post-epidemic threshold; C) Using high-intensity level. For both the pre-epidemic and post-epidemic thresholds, the silhouette method suggests 3 clusters, while 4 clusters are indicated for the high-intensity level.

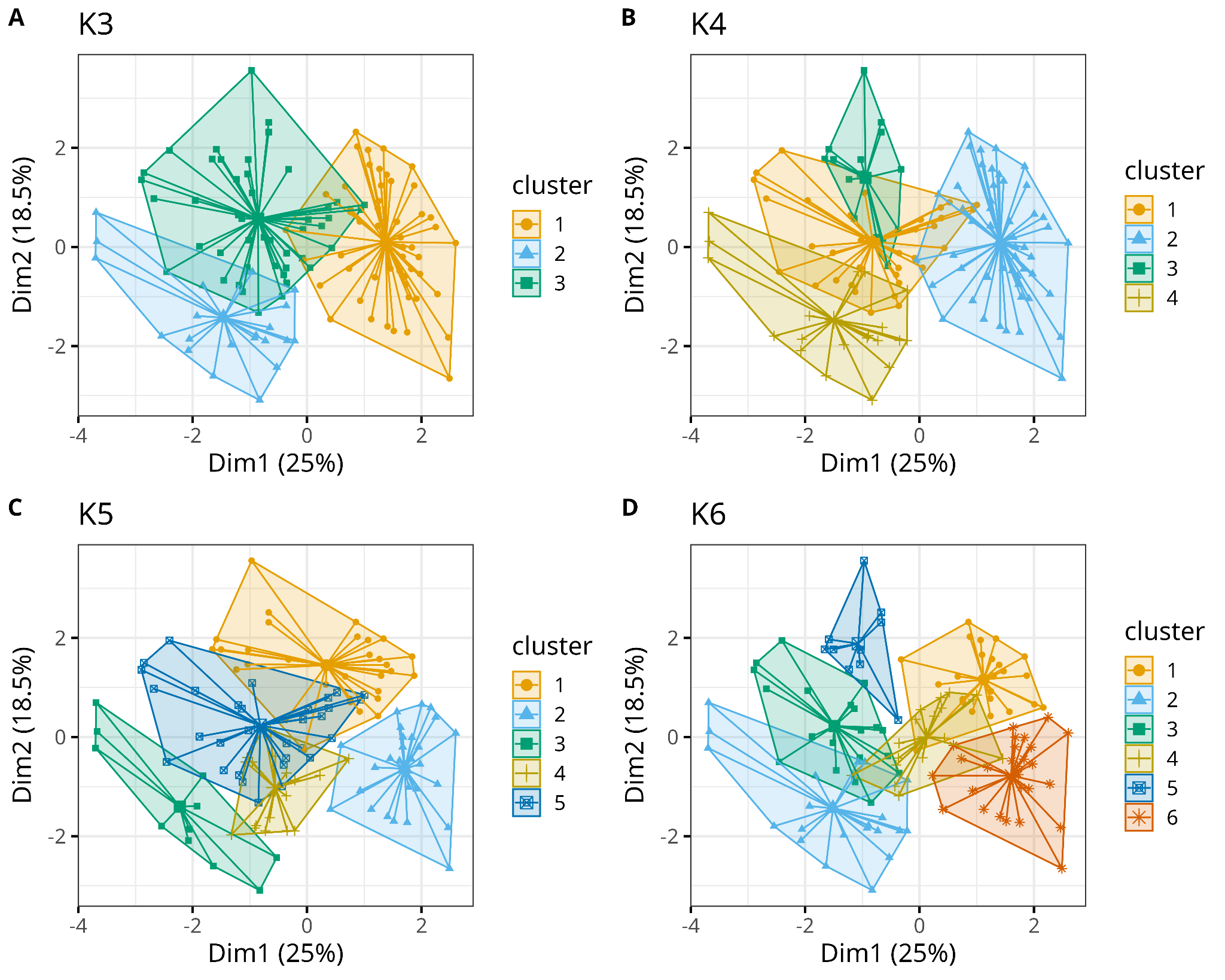

**S3-Figure 3.** Cluster plots using the pre-epidemic threshold. The K-means algorithm found similarities between several HMRs but it’s possible to note a lot of overlap between the clusters. A-D: 3-6 clusters.

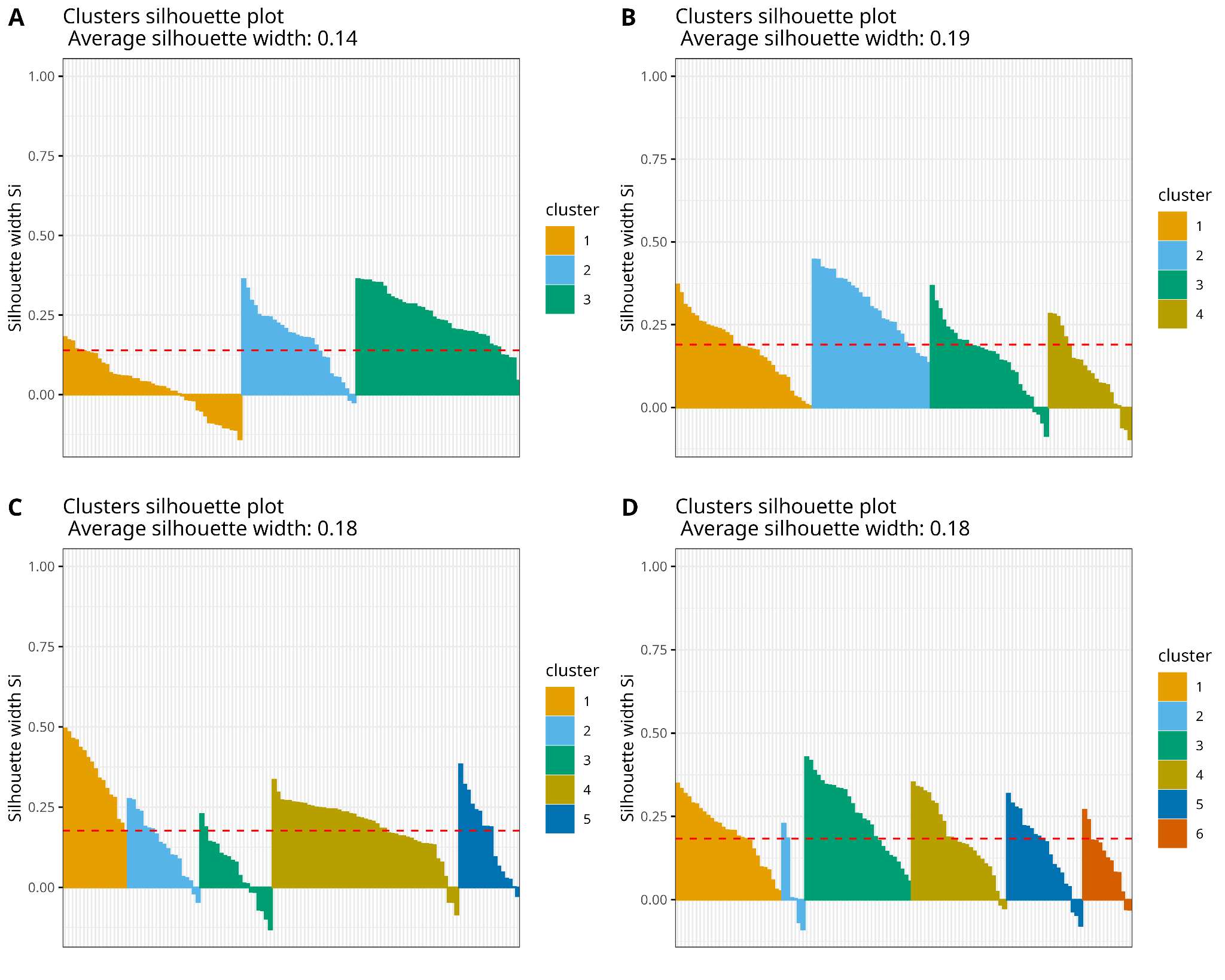

**S3-Figure 4.** Cluster quality using the pre-epidemic threshold and different numbers of clusters. A-D: 3-6 clusters. The increase in the number of clusters decreases the average silhouette indicating possible misclassifications.

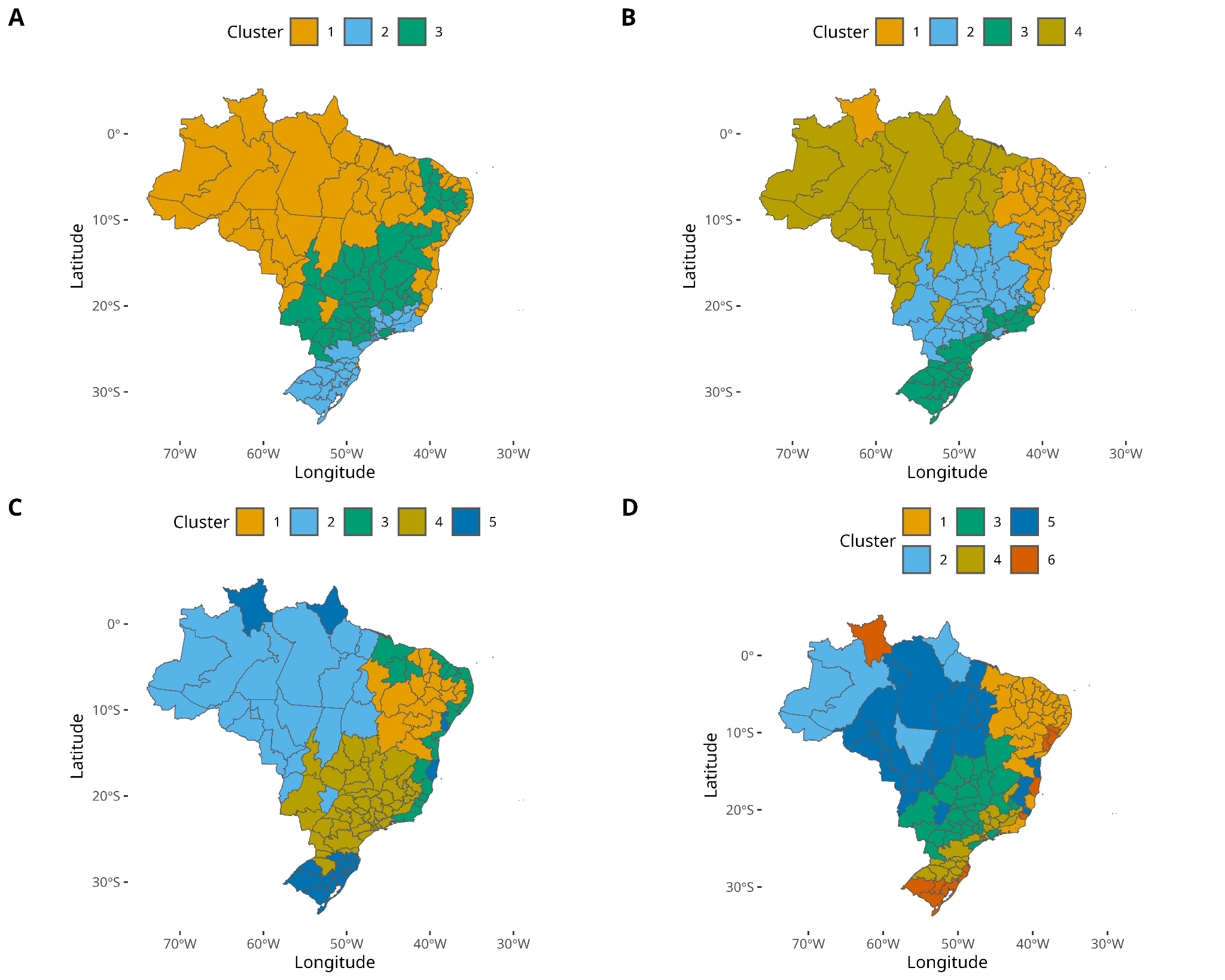

**S3-Figure 5.** Maps generated with the cluster analysis with pre-epidemic threshold. A-D: 3-6 Clusters. Service Layer Credits: Sources: <https://www.ibge.gov.br/geociencias/organizacao-do-territorio/malhas-territoriais/15774-malhas.html?=&t=downloads>

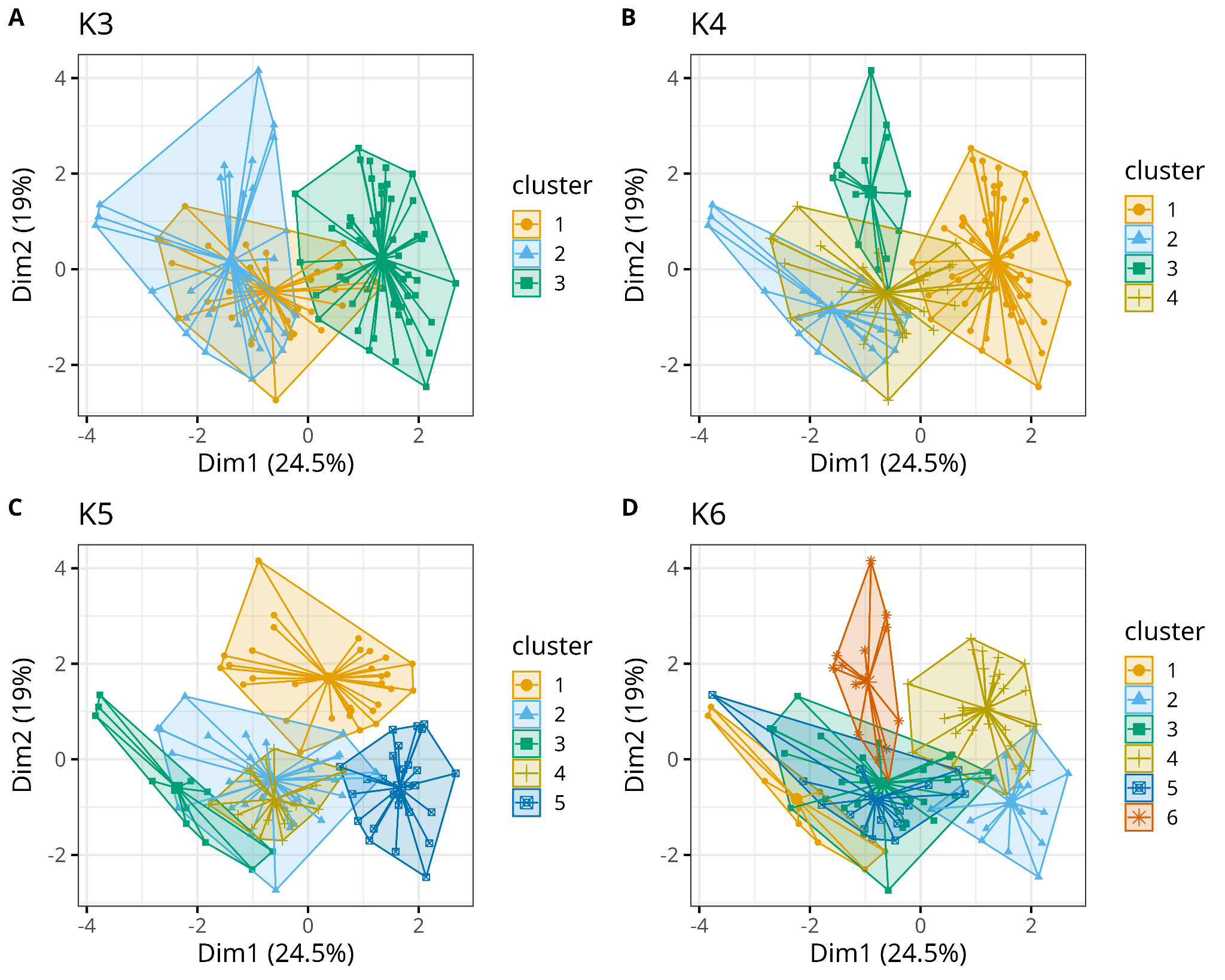

**S3-Figure 6**. Cluster plots using the post-epidemic threshold. The K-means algorithm found similarities between several HMRs but it’s possible to note a lot of overlap between the clusters. A-D: 3-6 clusters.

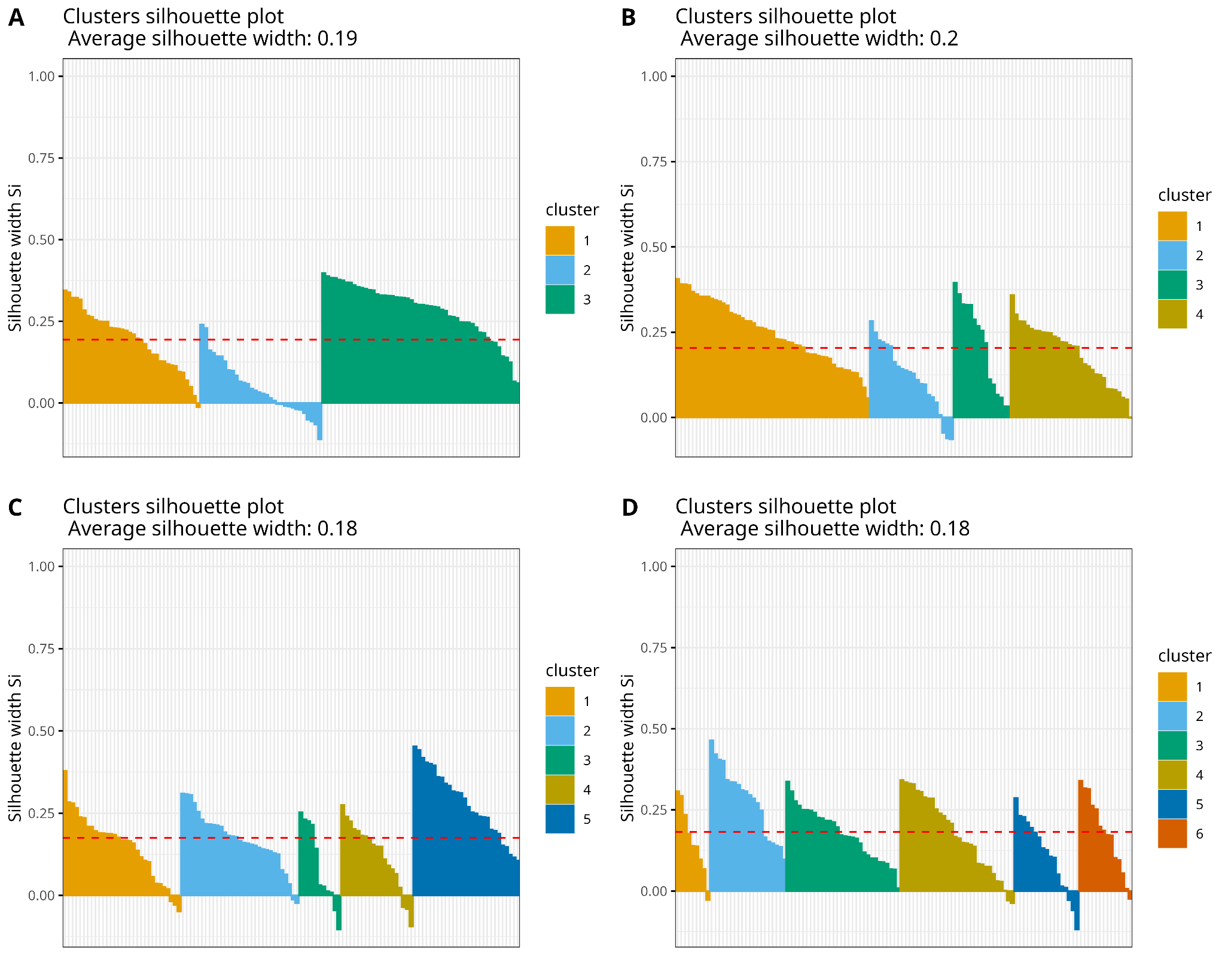

**S3-Figure 7.** Cluster quality using the post-epidemic threshold and different numbers of clusters. A-D: 3-6 clusters. The increase in the number of clusters decreases the average silhouette indicating possible misclassifications.

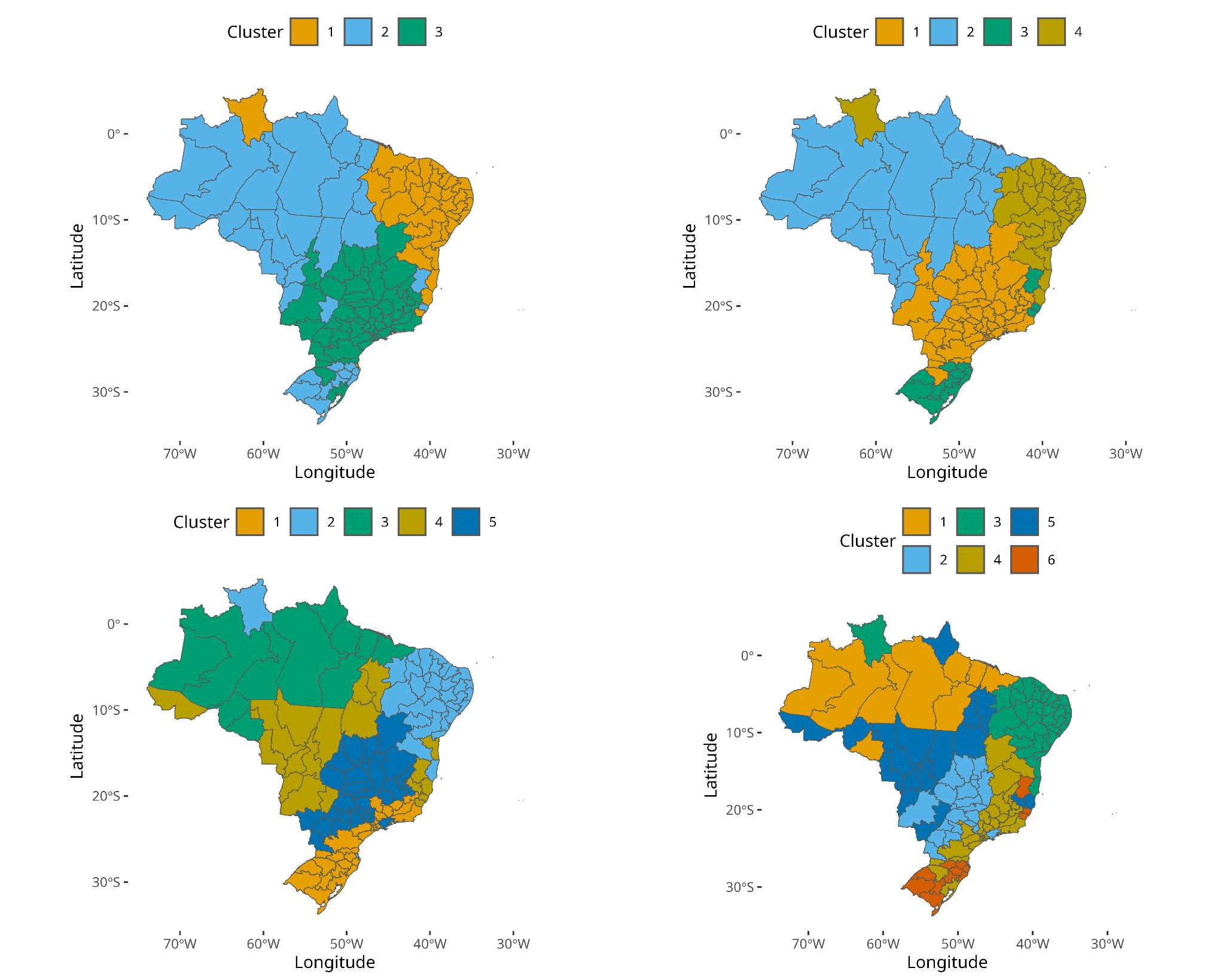

**S3-Figure 8.** Maps generated with the cluster analysis with pre-epidemic threshold. A-D: 3-6 Clusters. Service Layer Credits: Sources: <https://www.ibge.gov.br/geociencias/organizacao-do-territorio/malhas-territoriais/15774-malhas.html?=&t=downloads>

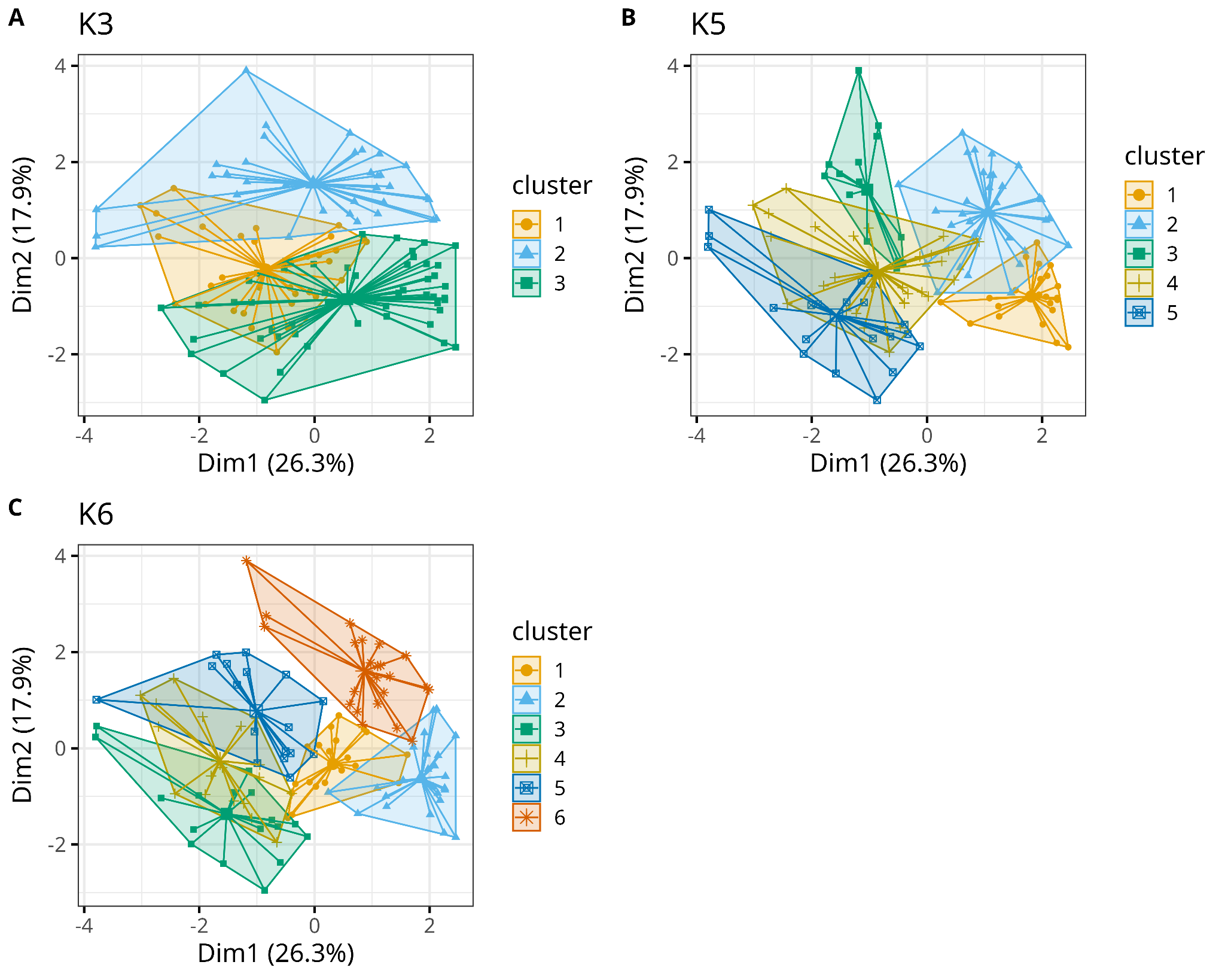

**S3-Figure 9**. Cluster plots using the high intensity level. The K-means algorithm found similarities between several HMRs but it’s possible to note a lot of overlap between the clusters. A-C: 3, 5 and 6 clusters. The plot with 4 clusters is in the main document.
