## Supplementary information 4 for "Unraveling Regional Variability in Dengue Outbreaks in Brazil: leveraging the Moving Epidemics Method (MEM) and Climate Data to Optimize Vector Control Strategies"

Dengue annual (2010-2023) incidence in each Health-Macro Region

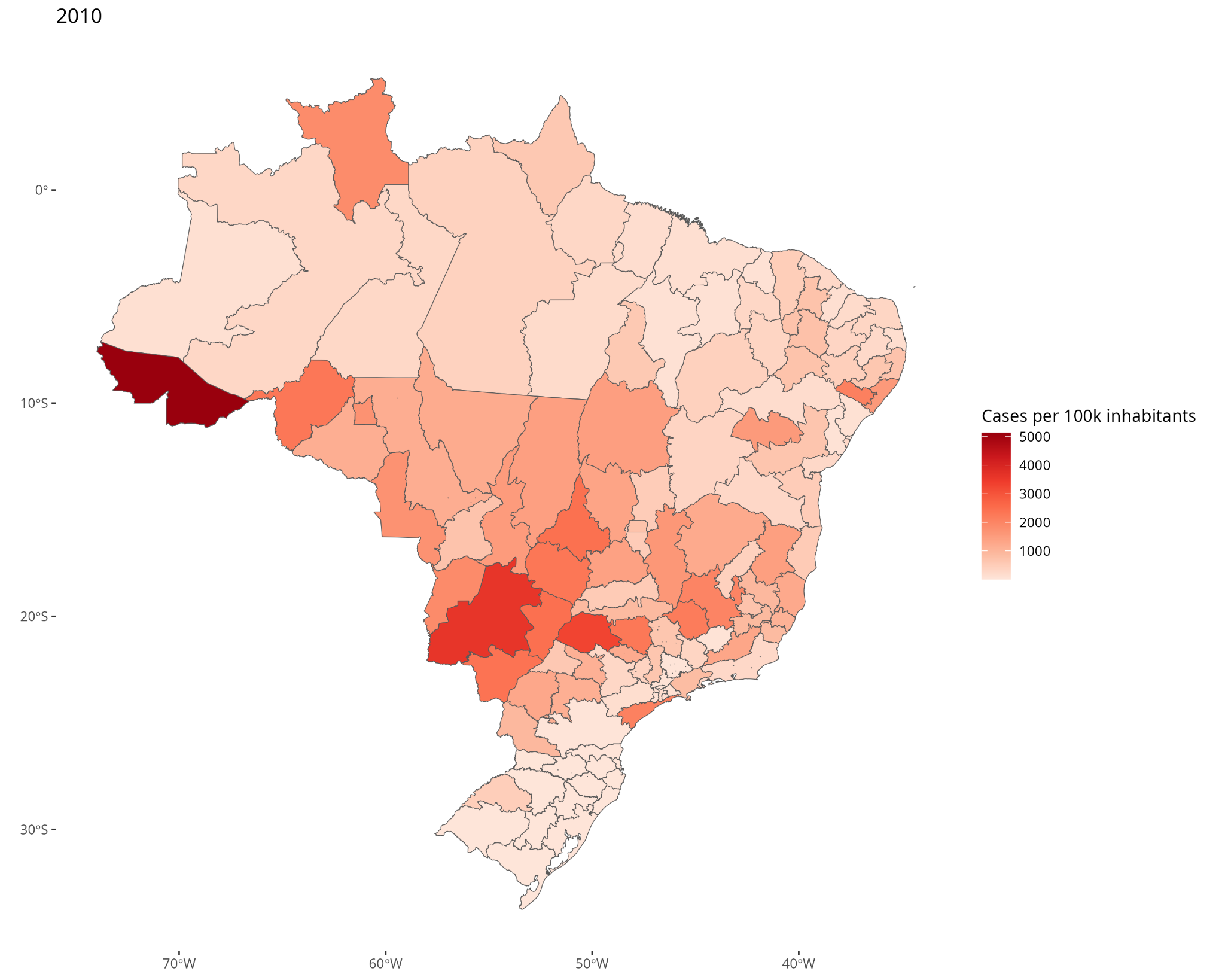

**S4 - Figure 1.** Number of dengue cases per 100,000 inhabitants for each Health-Macro Region (HMR) in 2010. Service Layer Credits: Sources:https://www.ibge.gov.br/geociencias/organizacao-do-territorio/malhas-territoriais/15774-malhas.html?=&t=downloads

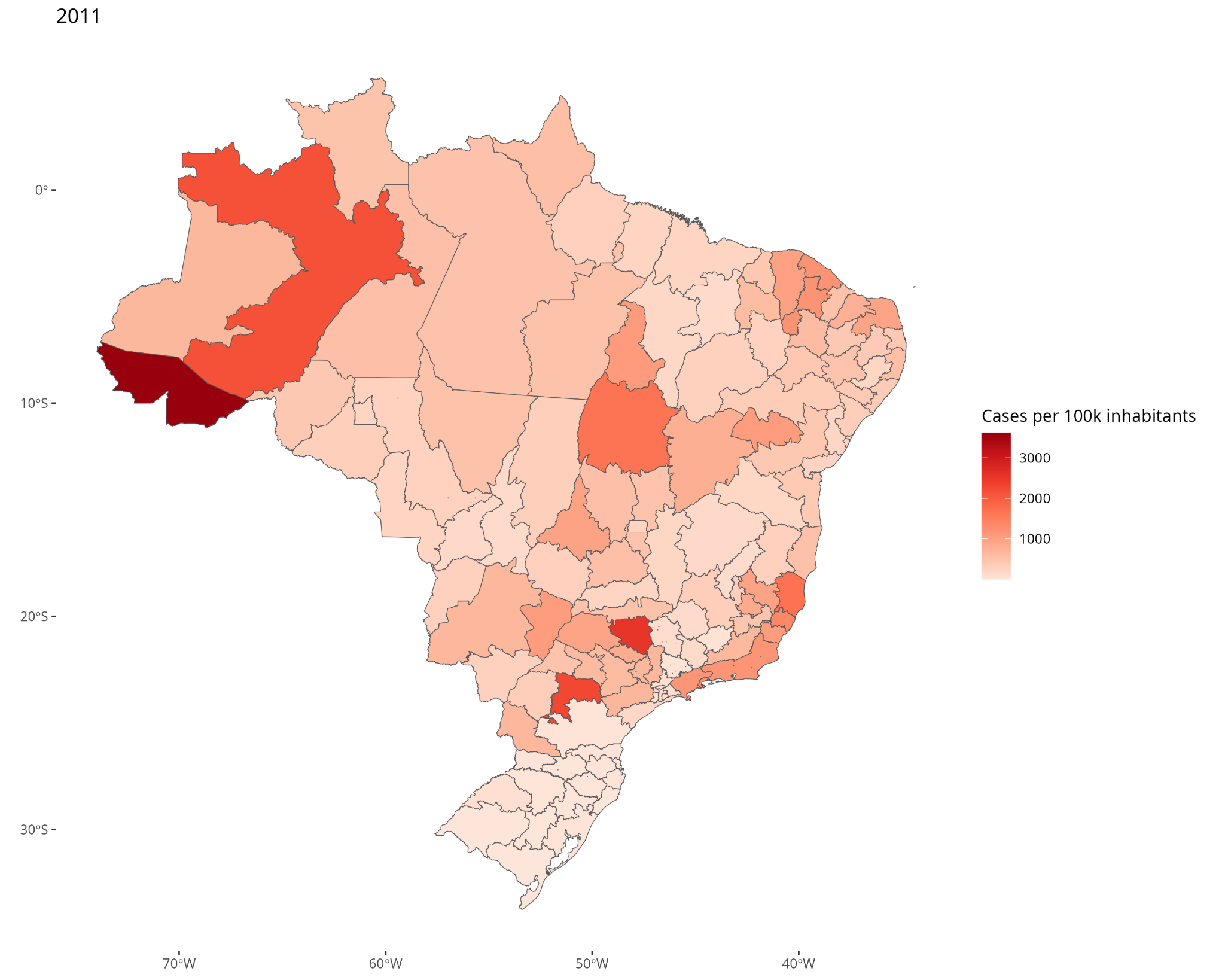

**S4 - Figure 2.** Number of dengue cases per 100,000 inhabitants for each Health-Macro Region (HMR) in 2011. Service Layer Credits: Sources:https://www.ibge.gov.br/geociencias/organizacao-do-territorio/malhas-territoriais/15774-malhas.html?=&t=downloads

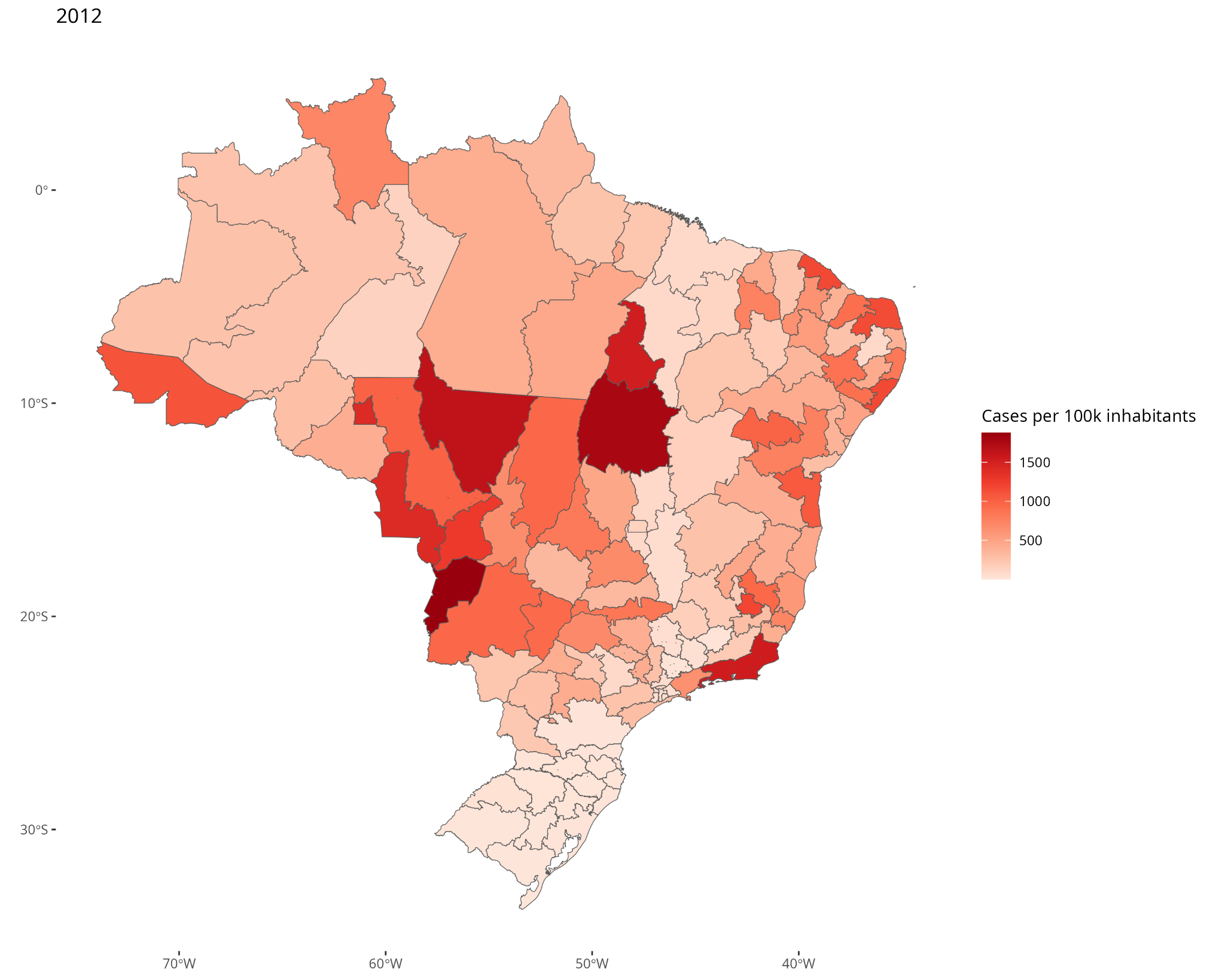

**S4 - Figure 3.** Number of dengue cases per 100,000 inhabitants for each Health-Macro Region (HMR) in 2012. Service Layer Credits: Sources:https://www.ibge.gov.br/geociencias/organizacao-do-territorio/malhas-territoriais/15774-malhas.html?=&t=downloads

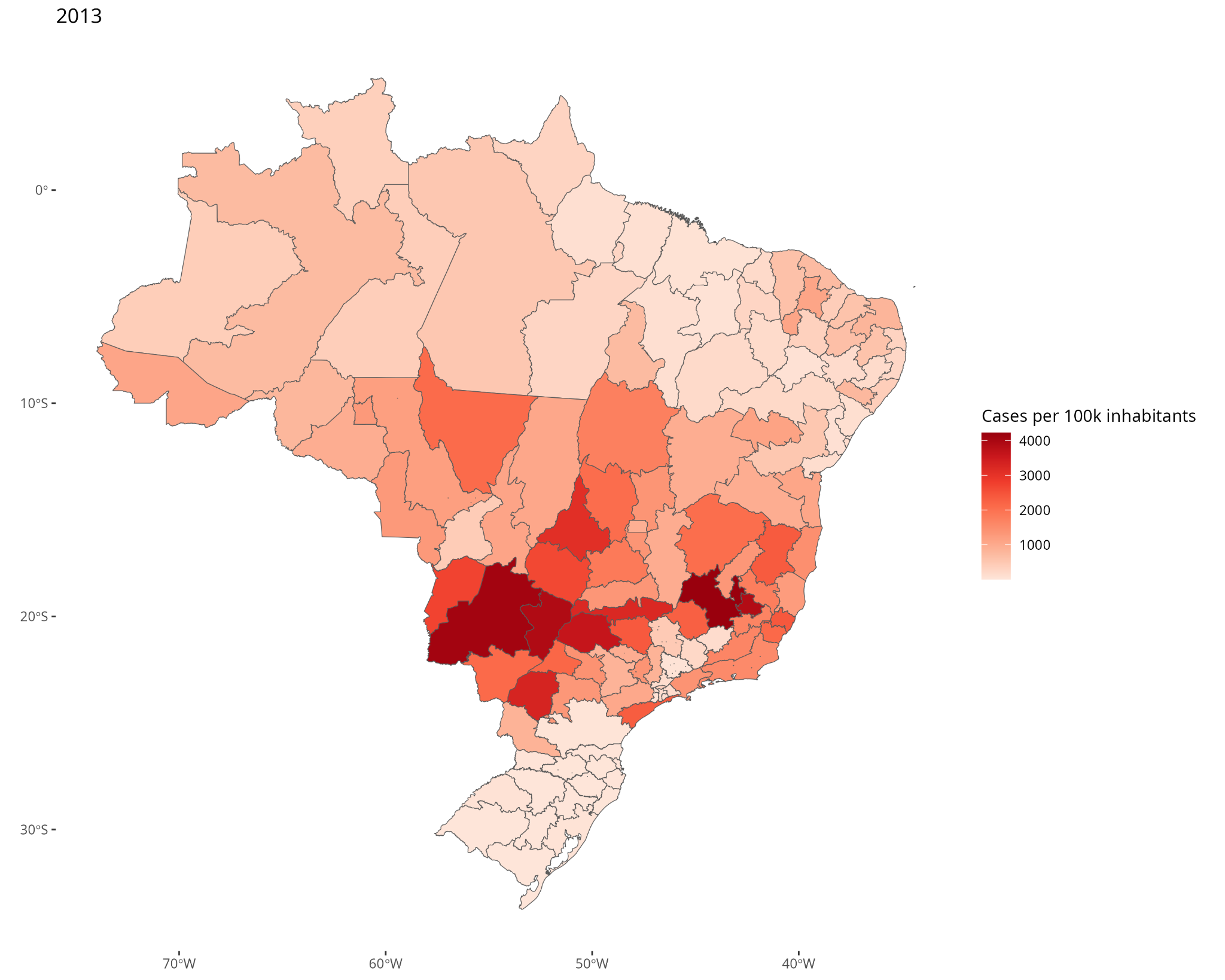

**S4 - Figure 4.** Number of dengue cases per 100,000 inhabitants for each Health-Macro Region (HMR) in 2013. Service Layer Credits: Sources:https://www.ibge.gov.br/geociencias/organizacao-do-territorio/malhas-territoriais/15774-malhas.html?=&t=downloads

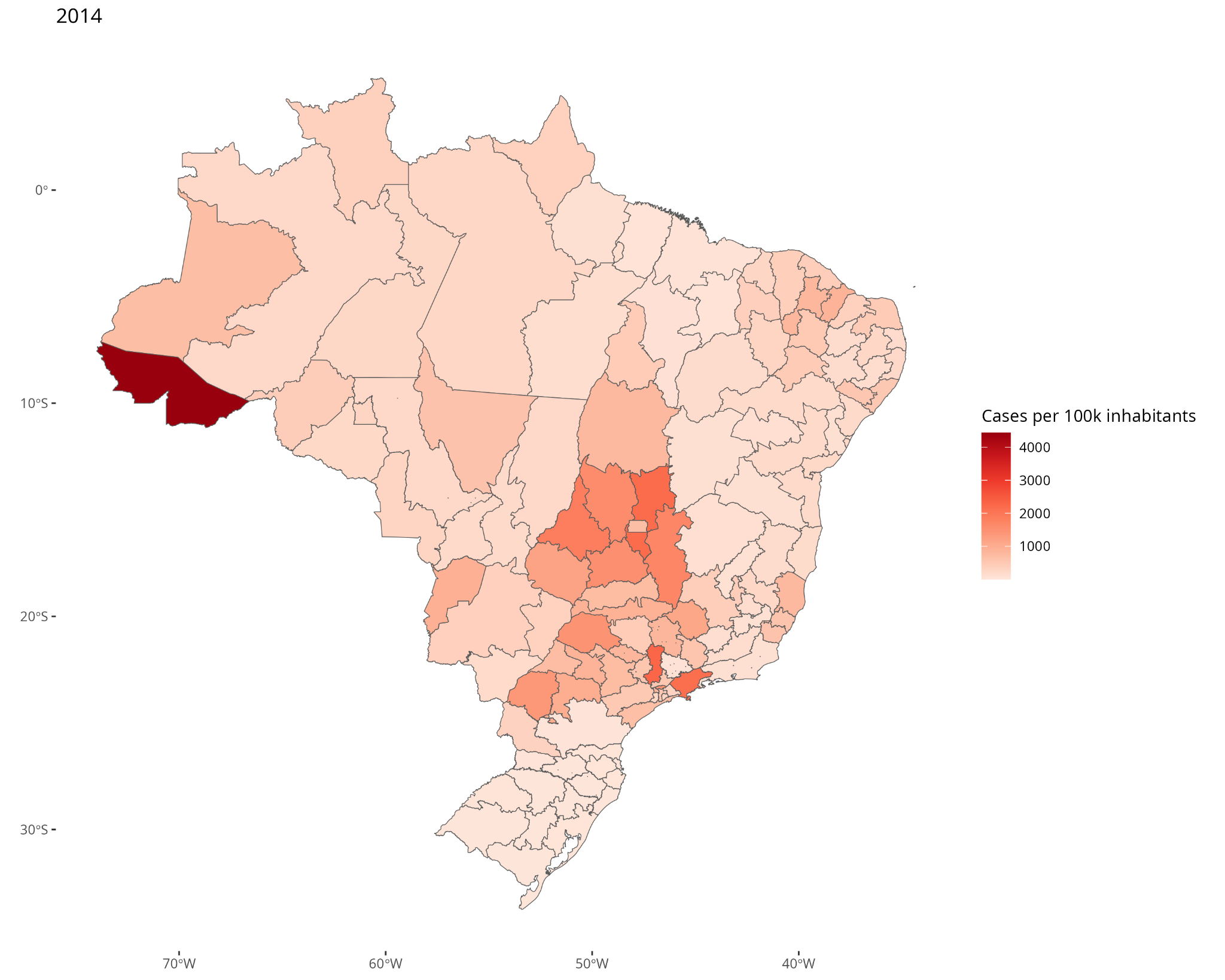

**S4 - Figure 5.** Number of dengue cases per 100,000 inhabitants for each Health-Macro Region (HMR) in 2014. Service Layer Credits: Sources:https://www.ibge.gov.br/geociencias/organizacao-do-territorio/malhas-territoriais/15774-malhas.html?=&t=downloads

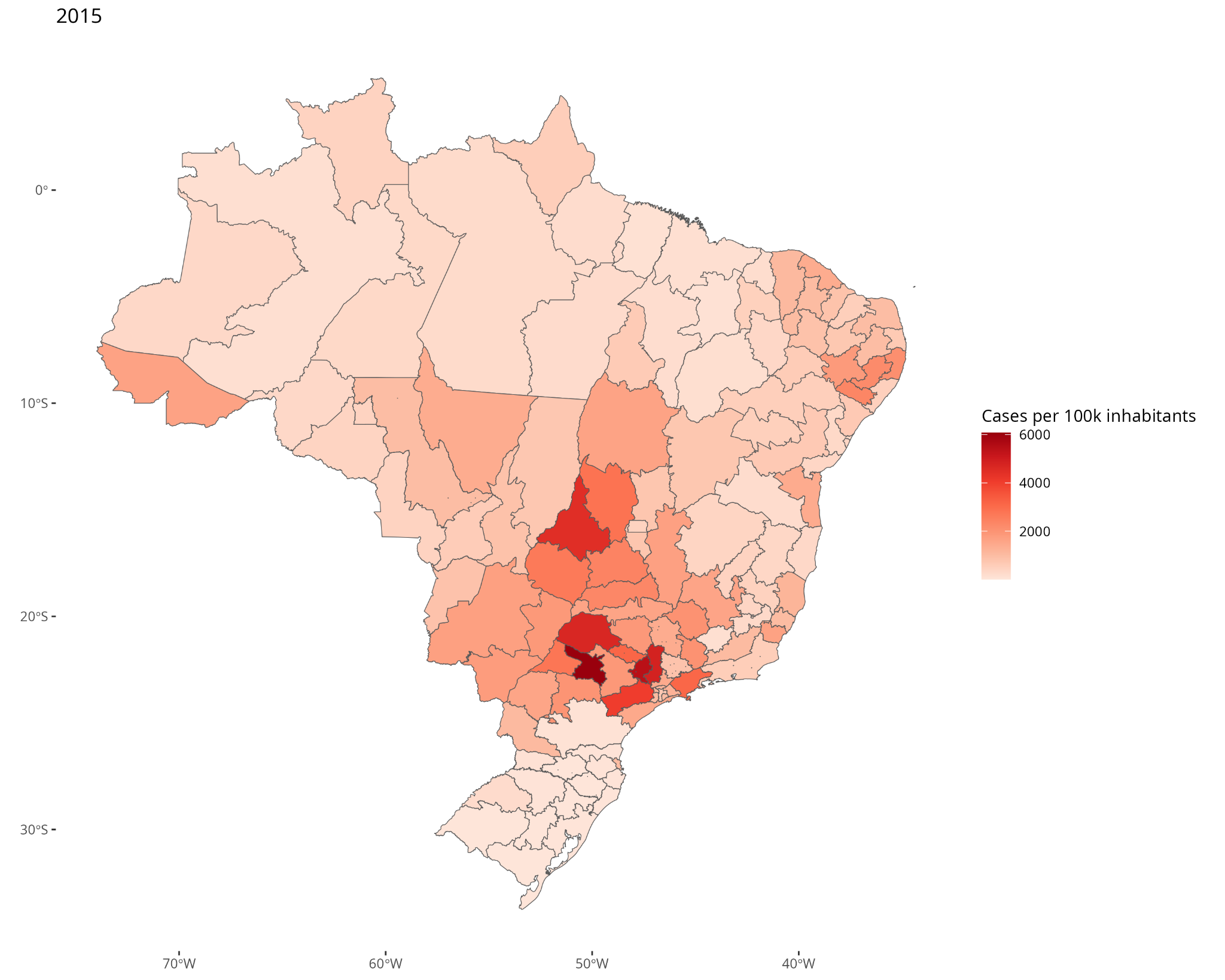

**S4 - Figure 6.** Number of dengue cases per 100,000 inhabitants for each Health-Macro Region (HMR) in 2015. Service Layer Credits: Sources:https://www.ibge.gov.br/geociencias/organizacao-do-territorio/malhas-territoriais/15774-malhas.html?=&t=downloads

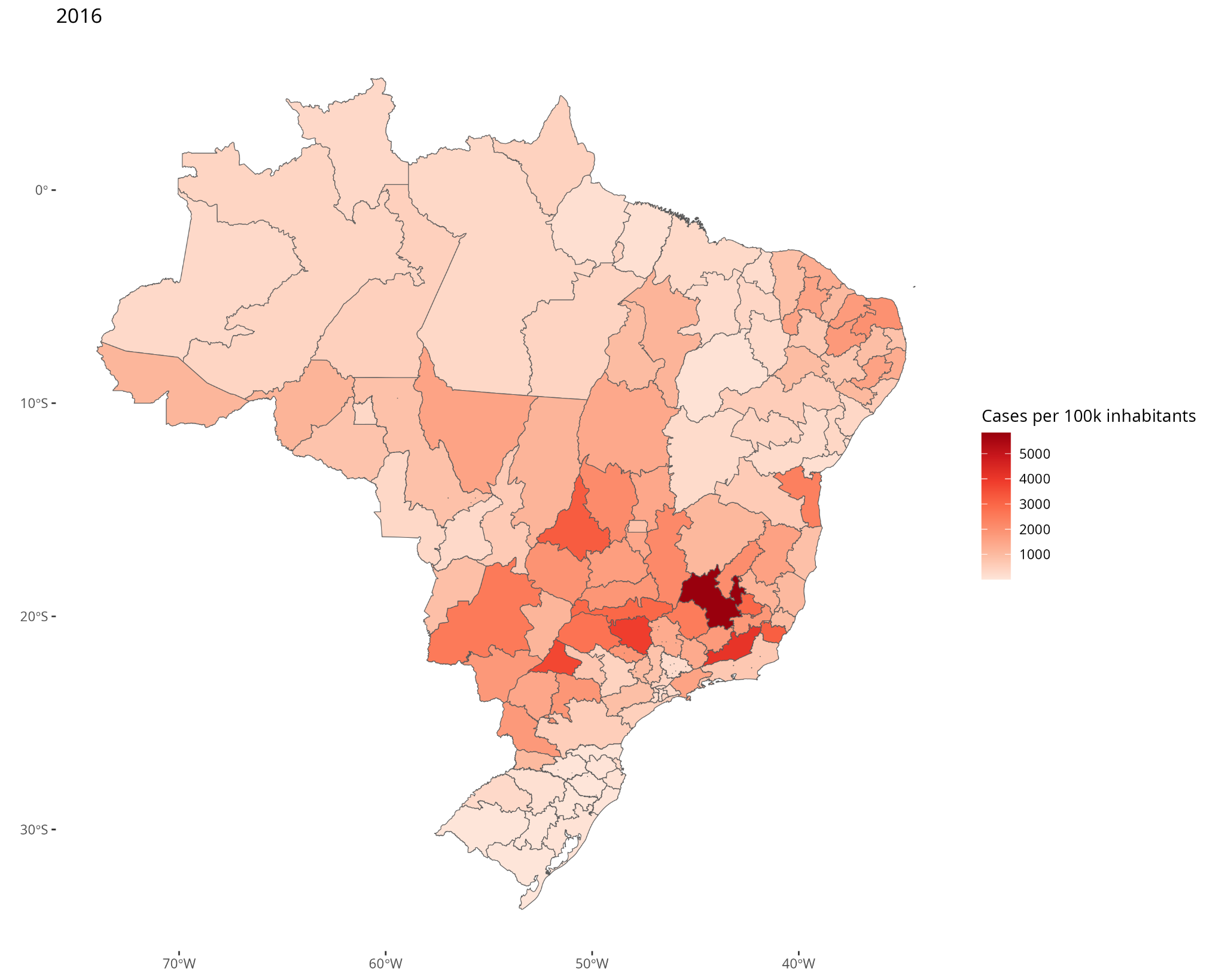

**S4 - Figure 7.** Number of dengue cases per 100,000 inhabitants for each Health-Macro Region (HMR) in 2016. Service Layer Credits: Sources:https://www.ibge.gov.br/geociencias/organizacao-do-territorio/malhas-territoriais/15774-malhas.html?=&t=downloads

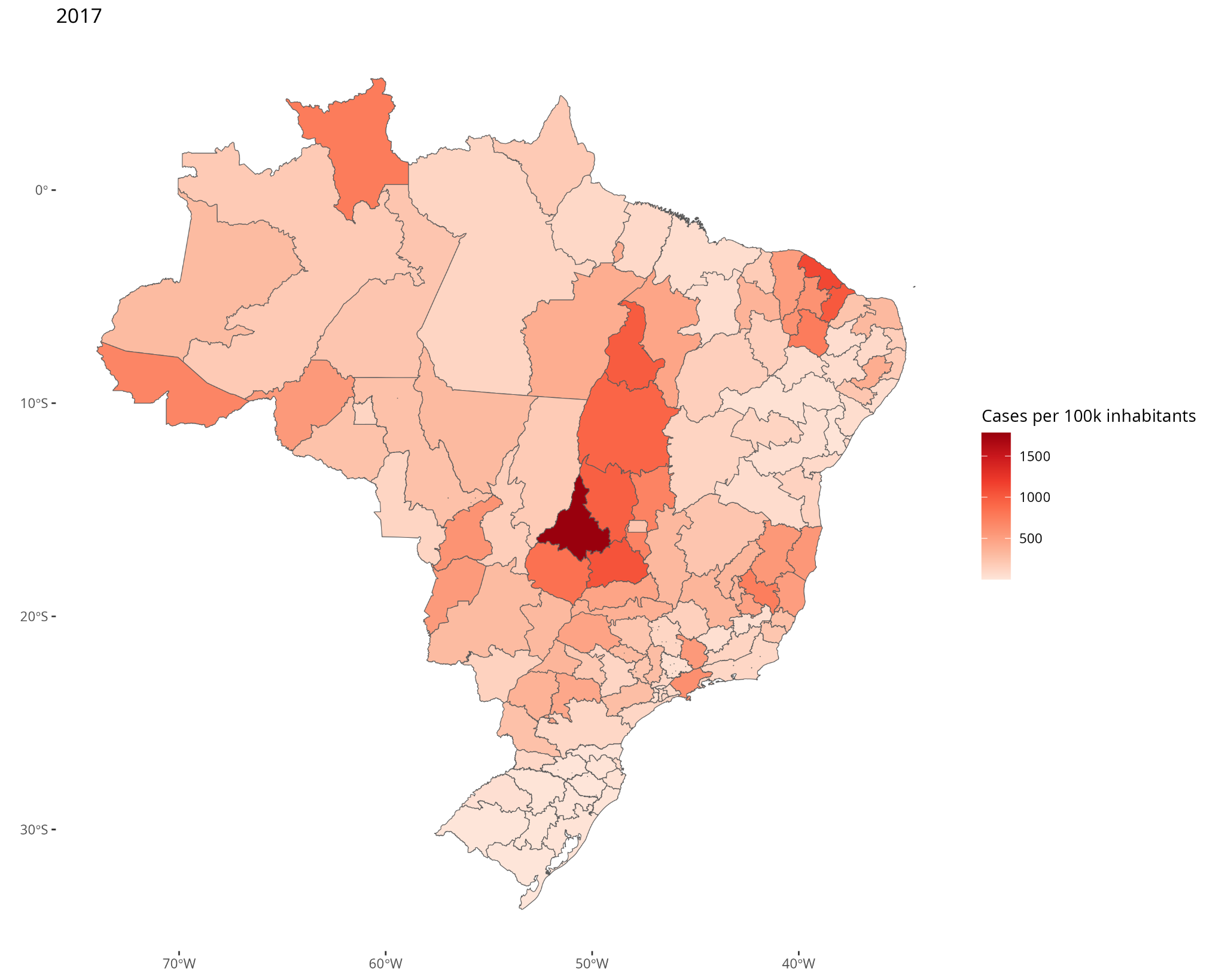

**S4 - Figure 8.** Number of dengue cases per 100,000 inhabitants for each Health-Macro Region (HMR) in 2017. Service Layer Credits: Sources:https://www.ibge.gov.br/geociencias/organizacao-do-territorio/malhas-territoriais/15774-malhas.html?=&t=downloads

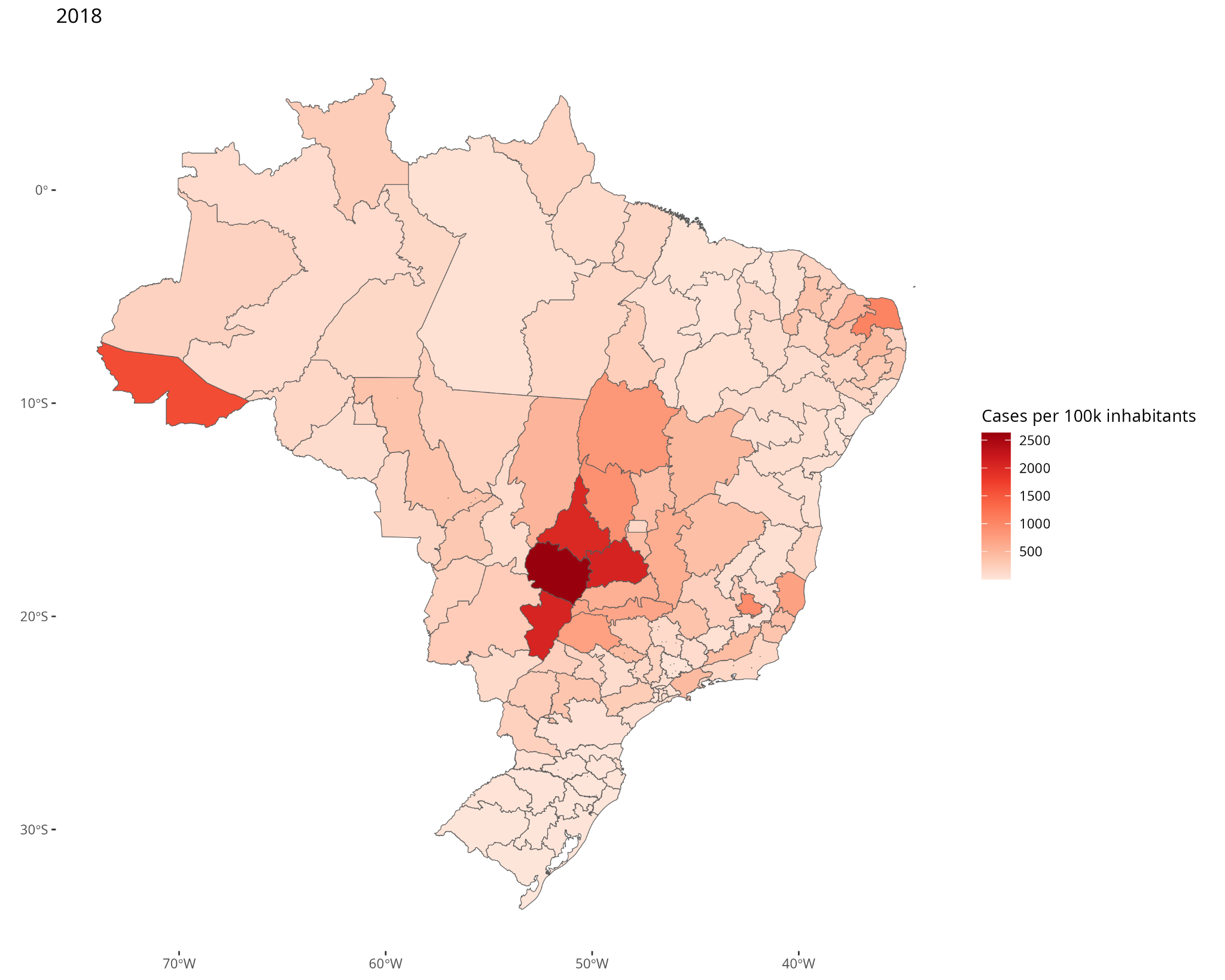

**S4 - Figure 9.** Number of dengue cases per 100,000 inhabitants for each Health-Macro Region (HMR) in 2018. Service Layer Credits: Sources:https://www.ibge.gov.br/geociencias/organizacao-do-territorio/malhas-territoriais/15774-malhas.html?=&t=downloads

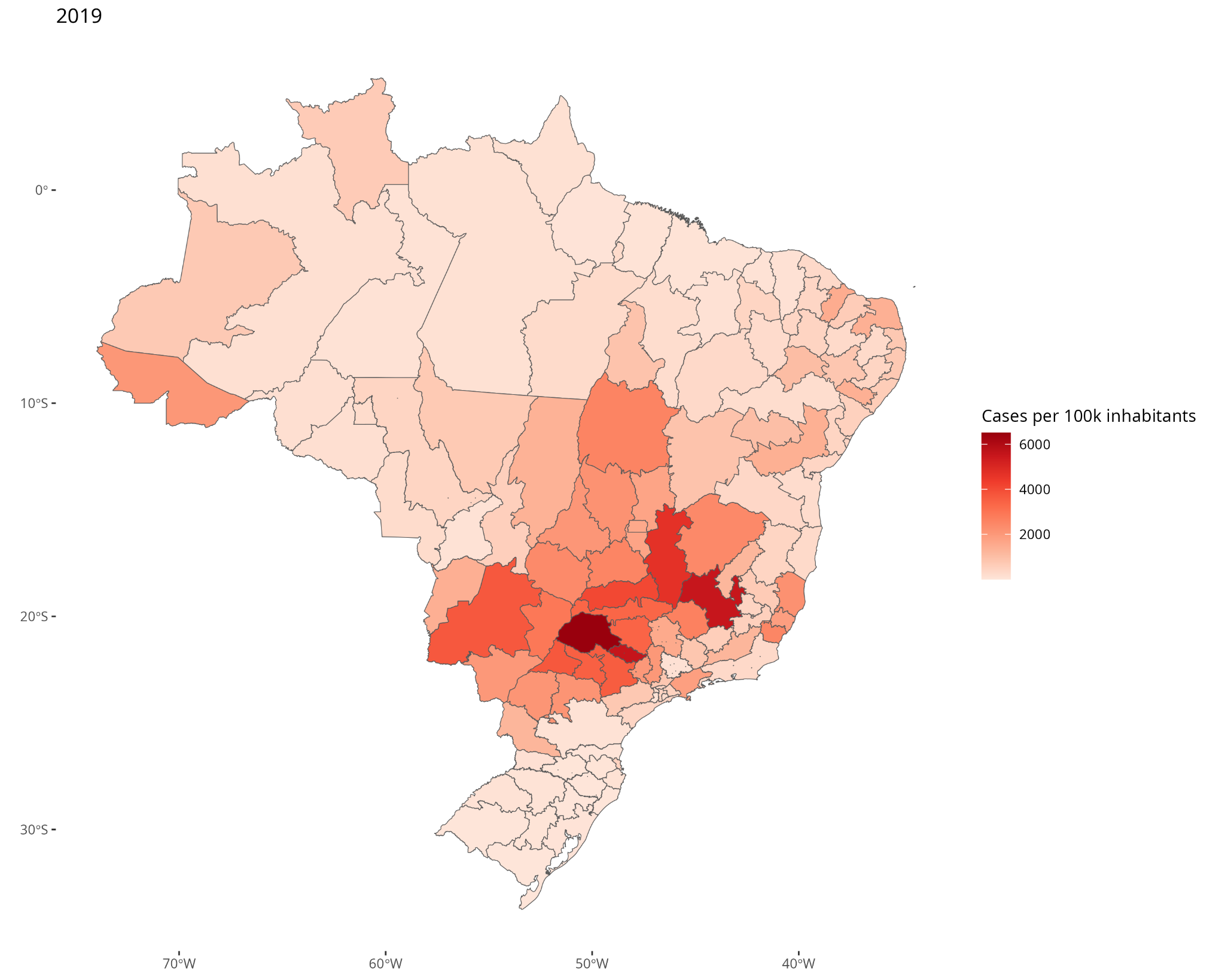

**S4 - Figure 10.** Number of dengue cases per 100,000 inhabitants for each Health-Macro Region (HMR) in 2019. Service Layer Credits: Sources:https://www.ibge.gov.br/geociencias/organizacao-do-territorio/malhas-territoriais/15774-malhas.html?=&t=downloads

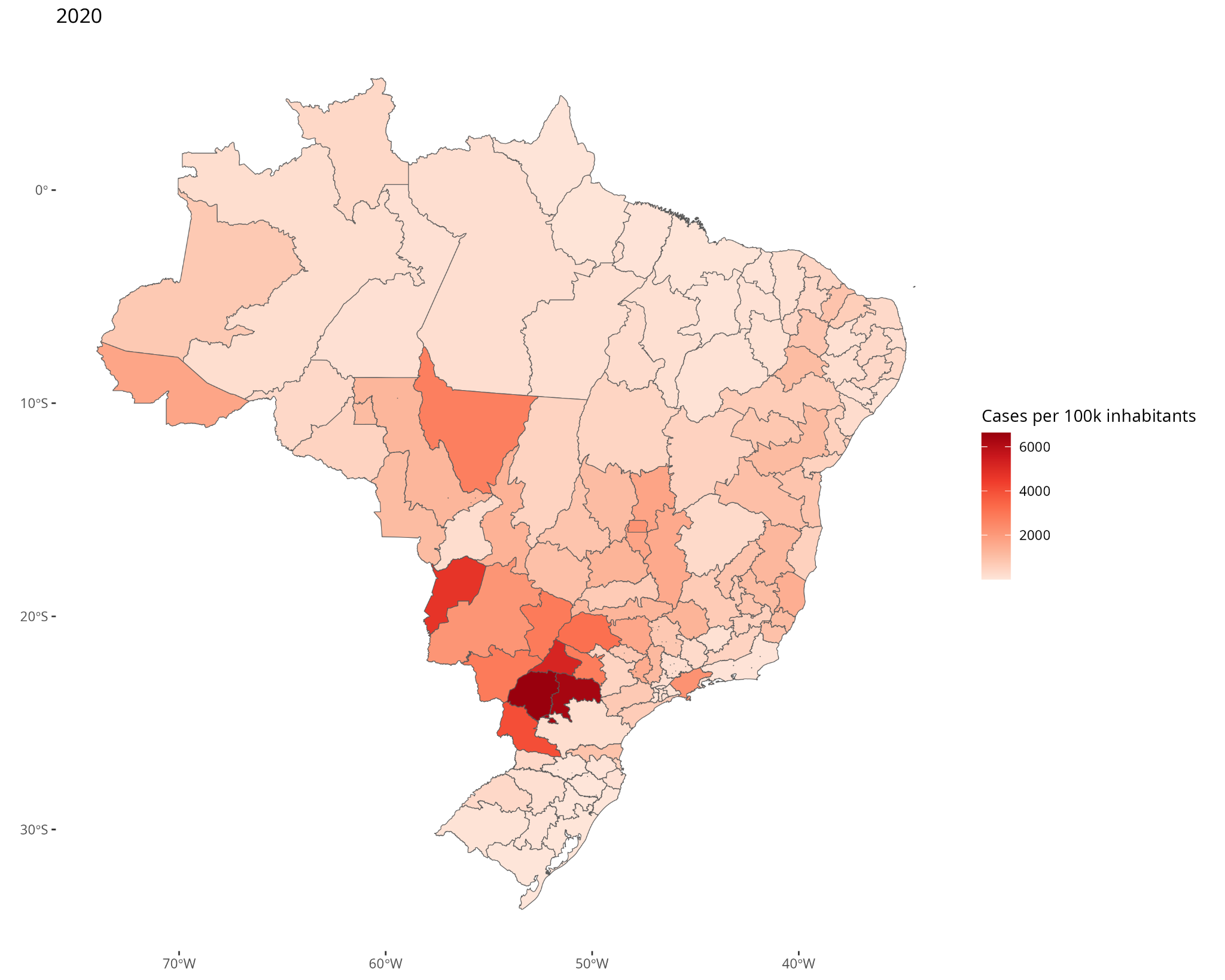

**S4 - Figure 11.** Number of dengue cases per 100,000 inhabitants for each Health-Macro Region (HMR) in 2020. Service Layer Credits: Sources:https://www.ibge.gov.br/geociencias/organizacao-do-territorio/malhas-territoriais/15774-malhas.html?=&t=downloads

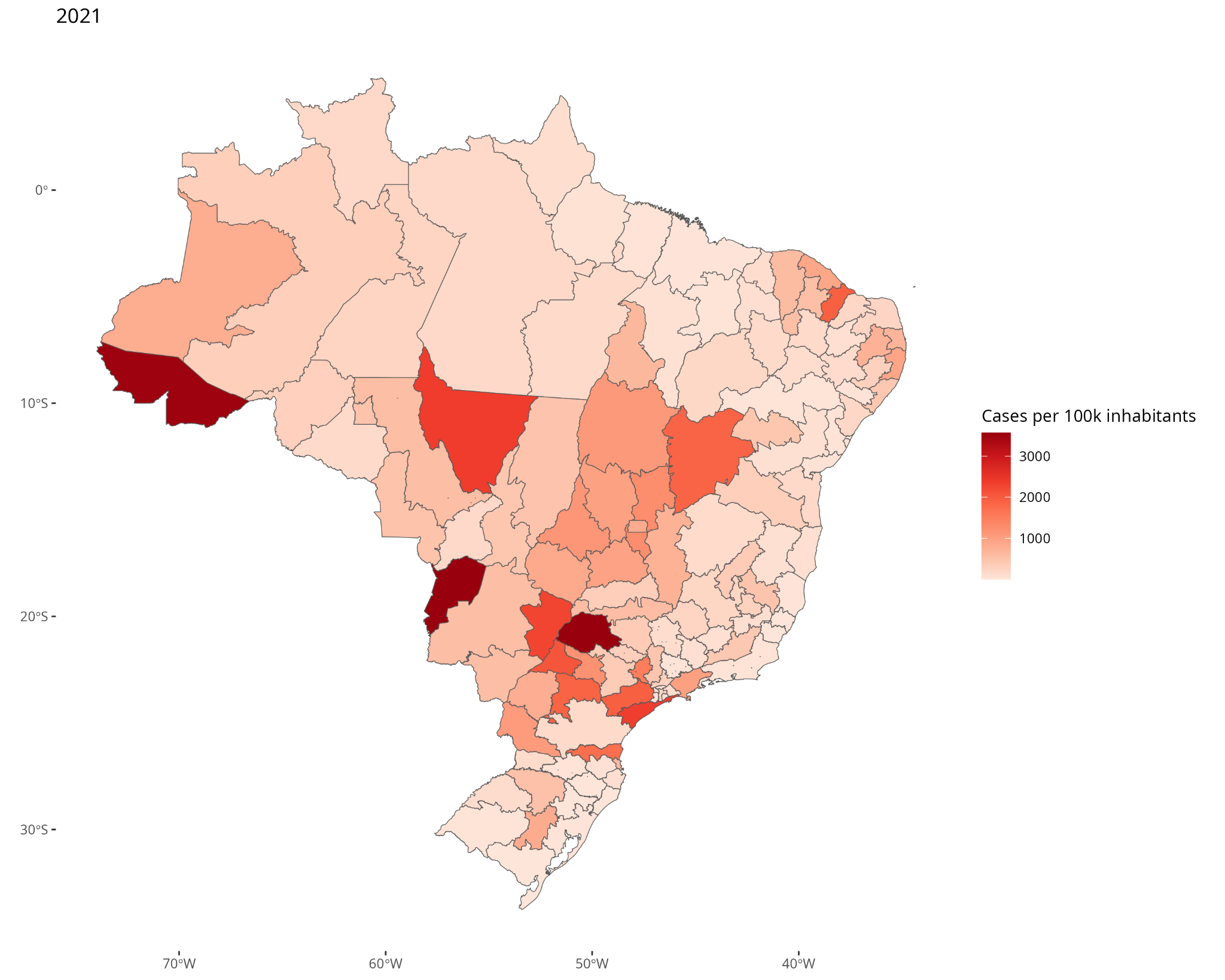

**S4 - Figure 12.** Number of dengue cases per 100,000 inhabitants for each Health-Macro Region (HMR) in 2021. Service Layer Credits: Sources:https://www.ibge.gov.br/geociencias/organizacao-do-territorio/malhas-territoriais/15774-malhas.html?=&t=downloads

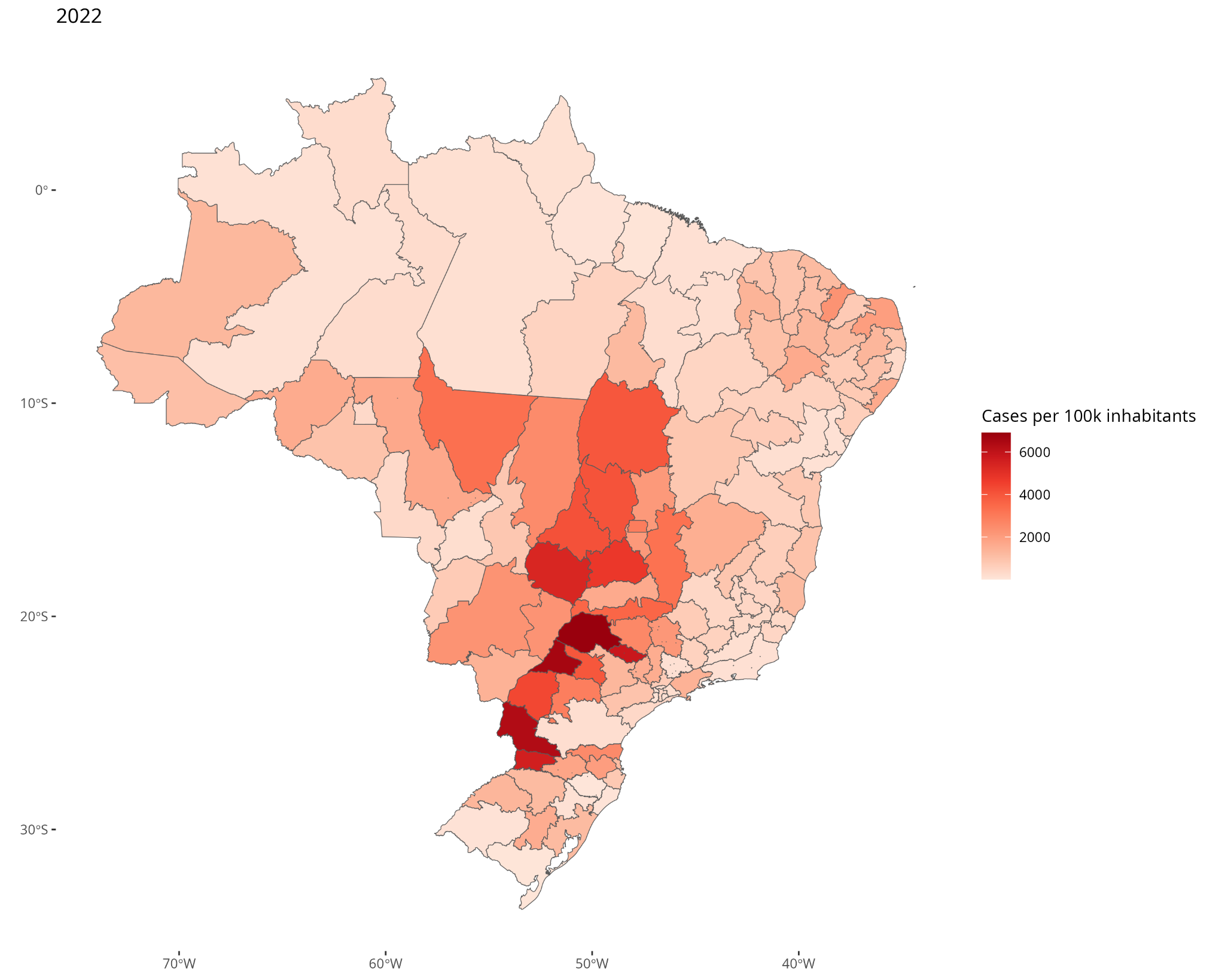

**S4 - Figure 13.** Number of dengue cases per 100,000 inhabitants for each Health-Macro Region (HMR) in 2022. Service Layer Credits: Sources:https://www.ibge.gov.br/geociencias/organizacao-do-territorio/malhas-territoriais/15774-malhas.html?=&t=downloads

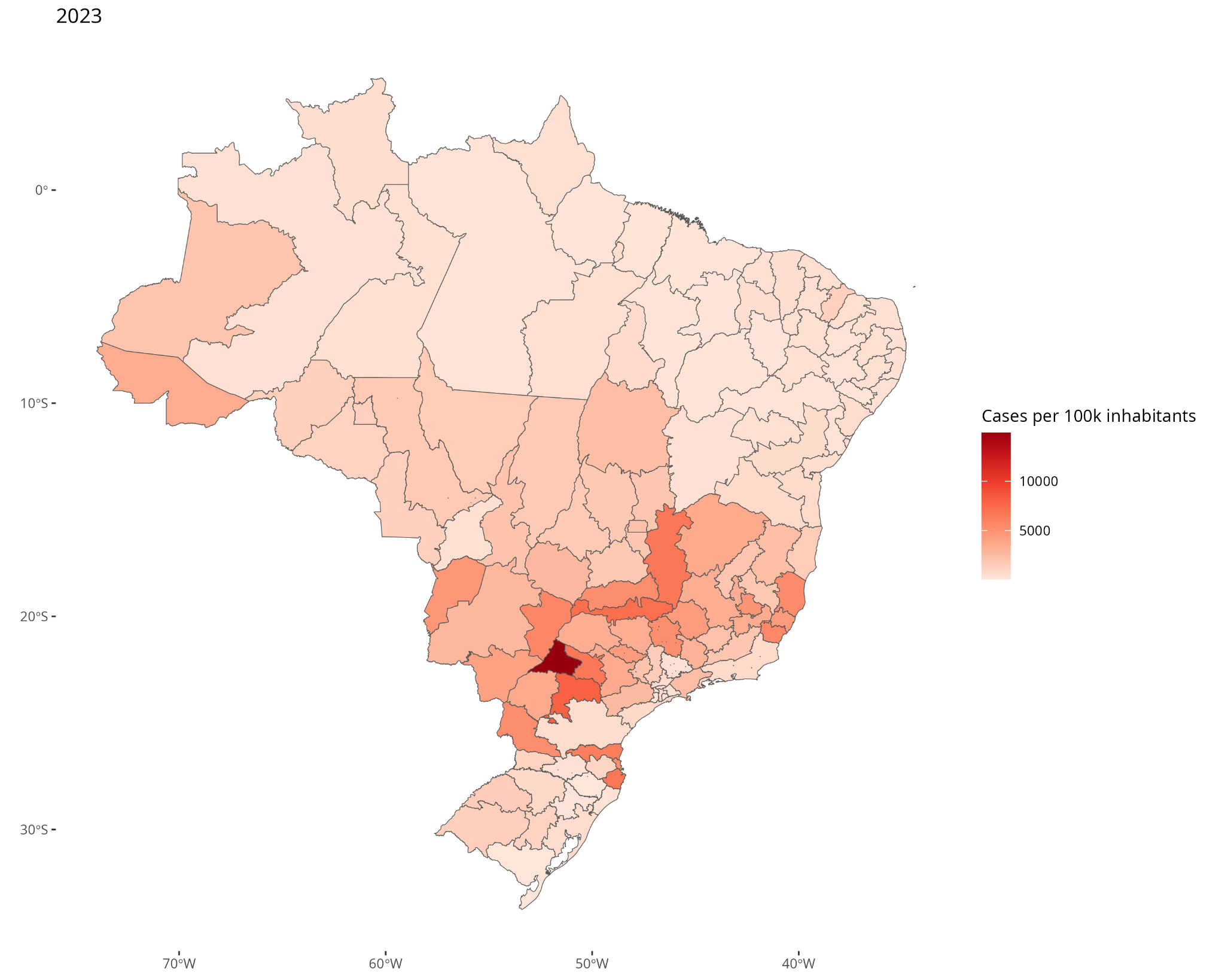

**S4 - Figure 14.** Number of dengue cases per 100,000 inhabitants for each Health-Macro Region (HMR) in 2023. Service Layer Credits: Sources:https://www.ibge.gov.br/geociencias/organizacao-do-territorio/malhas-territoriais/15774-malhas.html?=&t=downloads
